# Non-muscle Invasive Bladder Cancer Molecular Subtypes Predict Differential Response to Intravesical Bacillus Calmette-Guérin

**DOI:** 10.1101/2021.11.30.21266988

**Authors:** Florus C. de Jong, Teemu D. Laajala, Robert F. Hoedemaeker, Kimberley R. Jordan, Angelique C.J. van der Made, Egbert R. Boevé, Deric K.E. van der Schoot, Bart Nieuwkamer, Emiel A.M. Janssen, Tokameh Mahmoudi, Joost L. Boormans, Dan Theodorescu, James C. Costello, Tahlita C.M. Zuiverloon

## Abstract

The recommended treatment for patients with high-risk non-muscle invasive bladder cancer (HR-NMIBC) is tumor resection followed by adjuvant *Bacillus Calmette-Guérin* (BCG) bladder instillations. However, only 50% of patients benefit from this therapy. In case of progression to advanced disease, patients must undergo a radical cystectomy with significant morbidity and have a poor clinical outcome. Identifying tumors least likely to respond to BCG can translate into alternative treatments, such as early radical cystectomy or novel targeted or immunotherapies. Here we present molecular profiling of 132 BCG-naive, HR-NMIBC patients, and 44 post-BCG recurrences (34 matched), which uncovered three distinct BCG Response Subtypes (BRS1-3). Patients with BRS3 tumors have reduced recurrence and progression-free survival compared to BRS1-2. BRS3 tumors expressed high EMT-basal markers and had an immunosuppresive profile, which was confirmed with spatial proteomics. Tumors which recurred post-BCG were enriched for BRS3. BRS stratification was validated in a second cohort of 151 BCG-naive HR-NMIBC patients and the molecular subtypes outperformed guideline recommended risk stratification based on clinicopathological variables. For clinical application, we validated that a commercially approved assay was able to accurately predict BRS3 tumors (AUROC 0.86). Our findings provide a potential clinical tool for improved identification of HR-NMIBC patients at the highest risk of progression, which can be used to select patients for early radical cystectomy or novel subtype-directed therapies.

**One Sentence Summary:** Molecular subtypes are predictive of response to intravesical Bacillus Calmette-Guérin immunotherapy in non-muscle invasive bladder cancer.

## Introduction

Non-muscle invasive bladder cancer (NMIBC) accounts for 75% of all bladder tumors [1]. The recommended treatment for high-risk NMIBC (HR-NMIBC) consists of a transurethral resection of the bladder tumor (TURBT) and adjuvant intravesical *Bacillus Calmette-Guérin* (BCG) instillations [1]. Initial treatment response to BCG is excellent, yet the long-term efficacy is moderate as HR-NMIBC patients have a 50% risk of developing recurrent disease within 5 years [2]. Furthermore, a 20% risk of progression to advanced disease is observed, which is associated with high mortality [1, 3]. Patients with HR-NMIBC where a tumor recurs or progresses have been exposed to unnecessary BCG toxicity, a delay in radical treatment and have reduced survival [4, 5]. Therefore, it is critical to identify patients, who are at risk for treatment failure with standard-of-care, prior to initiation of BCG therapy [6, 7]. These patients might be candidates for an early radical cystectomy (RC) procedure, which although negatively affects quality of life, has excellent long-term outcomes [8]. Additionally, novel treatment modalities are urgently needed because of an ongoing global BCG shortage [9].

The genetic make-up of tumors can dictate therapeutic response, and it is hypothesized that molecular profiling of HR-NMIBC may reveal mechanisms of response to BCG [10, 11]. Several studies from whole-transcriptome analyses of NMIBC have led to clustering-based classification systems and identification of predictive signatures of disease progression [12-16]. *Dyrskjot et al*. prospectively validated a 12-gene expression signature that corresponded to progression in HR-NMIBC. However, the progression signature had limited added value compared to the standard clinicopathological risk stratification. Recently, a study identified five molecular subtypes in 73 HR-NMIBC patients treated with BCG, however none of the subtypes corresponded to progression-free survival [17]. A major drawback of NMIBC molecular subtyping studies has been the lack of BCG-treated patients, preventing the identification of specific molecular signatures predictive of BCG treatment response. Moreover, studies were hampered by a lack of detailed information on BCG treatment and a lack of patients who experienced progressive disease. In addition, a user-friendly tool for subtype identification has not yet been developed. Because of these caveats, the translational impact of molecular subtypes thus far remained limited.

In this study, we leverage two large and fully annotated cohorts of HR-NMIBC patients treated with BCG and with clinical follow-up to define three molecular subtypes that associate with clinical outcomes. The molecular subtypes have unique features that correlated with differential response to BCG. We identified an aggressive EMT-basal immunosuppressive subtype (BCG response subtype 3 (BRS3) and we show that BRS3 is the dominant subtype in post-BCG recurrences. We demonstrate that subtyping improved the guideline recommended risk stratification and that a commercially available pathway-assay was able to accurately identify BRS3 tumors for clinical utility. Finally, we identified druggable genes and pathways associated with resistance to immunotherapy in post-BCG recurrences as candidate targets for bladder-sparing therapies in HR-NMIBC patients.

## Results

### HR-NMIBC molecular subtypes with divergent clinical outcome after BCG

To identify gene signatures predictive of clinical outcome, we performed whole-transcriptome sequencing of two HR-NMIBC patient cohorts: discovery Cohort A, *n=*132 pre-BCG and *n=*44 post-BCG tumors (34 matched tumors) and validation Cohort B *n=*151 pre-BCG tumors. A synopsis of the study design is shown in **Fig. 1A**. Detailed information on resources and patient inclusion can be found in the methods section and all clinicopathological information and follow-up data for both cohorts is listed in **Table S1**. No differences in progression-free survival (PFS) were observed between Cohort A and B (**Fig. S1A**).

**Fig. 1.**
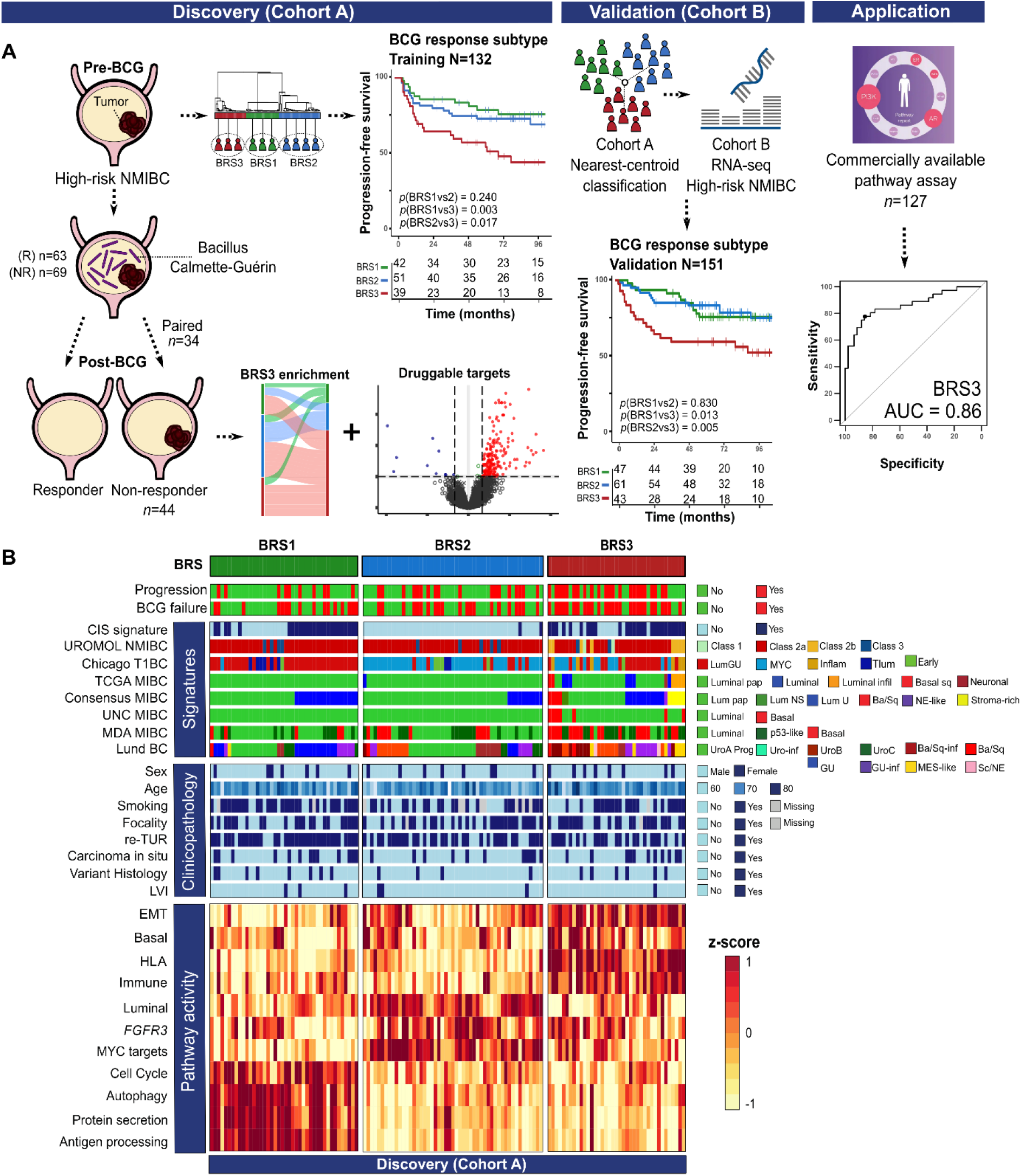
Study design, progression-free survival (PFS) and gene signatures of patients with high-risk NMIBC according to BCG response subtypes (BRS). **A:** RNA-seq was performed on Cohort A: *n*=132 pre-BCG, high-risk NMIBC tumors (*n*=63 BCG-responders [R] vs. *n*=69 BCG non-responders [NR]). From the BCG NR, *n*=44 post-BCG tumors were also sequenced (*n*=34 matched pre- and post BCG samples + *n=*10 non-matched). Paired analysis showed enrichment of BRS3 post-BCG and identified candidate druggable genes. Cohort B consisted of *n*=151 pre-BCG, high-risk NMIBC tumors (*n*=88 BCG-responders vs. *n*=63 BCG non-responders). For both cohorts, PFS is stratified according to BRSs. Finally, a qPCR pathway assay had an AUC of 0.86 in identifying BRS3 patients. **B:** Heatmap of gene signatures and annotation of Cohort A grouped according to BRS. From top to bottom: 1) Progression to MIBC; 2) BCG response; 3) carcinoma in situ signature [19];4) UROMOL21 NMIBC subtypes [13]; 5) T1BC subtypes [17]; 6) TCGA MIBC subtypes [18]; 7) Consensus MIBC subtypes [17]; 8) UNC MIBC subtypes [20]; 9) MDA MIBC subtypes [21]; 10) Lund BC subtypes [14]; 11) Clinicopathological parameters associated to pre-BCG tumors; 12) BRS signatures based on mean gene expression of selected genes (details in methods). **Abbreviations**: BCG = Bacillus Calmette-Guérin; (N)MIBC = (non-)muscle-invasive bladder cancer.

To determine the presence and robustness of molecular subtypes, we split Cohort A at a 3:1 ratio into a training and testing set. We applied consensus clustering on the top 2000 protein coding genes with the most varying expression in the training set to identify three subtypes with differential risk of progression (**Fig. S1B-C**). To prevent bias caused by immune populations based on gene expression analyses, all clustering in this study was performed with removal of genes predictive of immune cell populations. A nearest shrunken centroid classifier was then trained and used to predict subtypes in the testing set. Based on estimated PFS and overlapping pathway activity, the presence of three molecular subtypes was confirmed in the testing set (**Fig. S1D-E**). Subtypes were called BCG Response Subtype 1, 2 and 3 (BRS). After cross validation to estimate the robustness of the BRS in the training and testing set, we retrained the model on the full dataset of Cohort A (**Fig. S1F)**, and set out to investigate the clinical and molecular features of the BRS. Patients with BRS3 tumors had a significantly worse PFS (**Fig. 1A**) than BRS1/2, with a 2-year PFS of 61% for BRS3 versus 78% for BRS2 and 83% for BRS1. Differential pathways associated with the BRS in Cohort A are illustrated in **Fig. 1B)**, the top 30 non-synonymous SNVs are in **Fig. S2** (**Table S2**). BRS3 patients showed increased EMT pathway activity, and were significantly enriched for mutations associated with the extracellular matrix (ECM) (e.g. *AEBP1*), which might explain the higher risk of progression in BRS3 patients.

Next, we set out to validate the BRSs clinical outcome and molecular features. To this end, we performed whole-transcriptome sequencing on a second HR-NMIBC patient cohort (Cohort B, *n=*151 pre-BCG tumors; see **Table S1** for patient information) and used the BRS classifier trained on Cohort A to predict molecular subtypes in Cohort B. **Fig. S3** illustrates highly similar pathway activity per BRS in patients from Cohort B vs A. Here too, survival analysis showed that patients with BRS3 tumors had the worst PFS (2-year PFS 94% for BRS1, 87% for BRS2 and 67% for BRS3, **Fig. 1A**). To explore how gene expression of the BRSs would overlap with previously published NMIBC and MIBC subtypes, we applied the BRS classifier to UROMOL21 and TCGA cohorts [13, 18]. In UROMOL21, BRS3 predicted tumors were overlapping with the more aggressive Class 2A/B NMIBC subtype, while the less aggressive Class 1/3 tumors were more likely to be BRS1/2 (*p*<0.01, **Fig. S4A**). Although UROMOL21 contained patients that were not primary HR-NMIBC or treated with ≥5 instillations of BCG, we did observe overlapping gene signatures and a similar BRS stratification, which was specifically evident for stage T1 HR-NMIBC (**Fig. S4B**). In the TCGA cohort, BRS1/2 tumors were predominantly luminal papillary (*p*<0.001). BRS3 tumors were more frequently luminal infiltrated and basal/squamous (*p*<0.001) (**Fig. S4C**), and these tumors are known to be more aggressive. As a final step, we validated pathway activity in a second independent dataset with BCG-treated, HR-NMIBC patients (**Fig. S4D**). BRS2 (MYC high) seemed to associate with T1-MYC, while BRS3 (basal high) did not contain any T1-Lum tumors. No association with PFS was observed, but only eight patients progressed [17].

Last, we investigated whether clinicopathological parameters could explain differences between subtypes in the combined dataset (**Table 1**), or in Cohort A or B separately **(Table S1A-B)**. Due to the probability of understaging in 10% of T1 HR-NMIBC and thus BCG treatment failure, guidelines recommend that patients undergo a re-TURBT prior to initiation of BCG treatment [22]. Although a re-TURBT itself was not associated with PFS (HR = 1.0, *p*=0.97), the proportion of patients who had undergone a re-TURBT was higher in BRS1 patients, who had a favorable outcome. We recognize that a difference in re-TURBTs might lead to a decreased risk of treatment failure. Therefore, we repeated clustering in only patients who received a re-TURBT 91/132 (69%). Neither risk stratification, nor expression patterns differed between patients with and without re-TURBT **(Fig. S5)**, thus we concluded that a re-TURBTs did not affect the prognostic capability of the BRS. In summary, we discovered three molecular subtypes (BRS) of HR-NMIBC and showed their clinical relevance and consistency in pathway activity across multiple patient cohorts.

**Table 1.** Baseline patient characteristics and clinical follow-up of all *n*=283 BCG-naive high-risk non-muscle invasive bladder cancer patients treated with BCG and stratified according to subtype.

| Patient characteristics <sup>a</sup> |  | All<br>*n* (%) | BRS1<br>*n* (%) | BRS2<br>*n* (%) | BRS3<br>*n* (%) | *p* |
| --- | --- | --- | --- | --- | --- | --- |
| Sex | male | 56 (80) | 70 (25) | 90 (33) | 67 (24) | 0.895 |
|  | female | 227 (20) | 19 (7) | 22 (8) | 19 (5) |  |
| Age | Median (IQR) | 70 (62-77) | 70 (62-77) | 69 (62-76) | 71 (62-77) | 0.826 |
| Smoking | No | 87 (31) | 23 (8) | 36 (13) | 28 (10) | 0.432 |
|  | Yes/stopped | 173 (61) | 61 (22) | 64 (23) | 64 (17) |  |
|  | Missing | 23 (8) | 5 (2) | 12 (4) | 6 (2) |  |
| re-TURBT | No | 93 (32) | 23 (8) | 37 (13) | 33 (12) | 0.138 |
|  | Yes | 190 (67) | 66 (23) | 75 (27) | 49 (17) |  |
| Focality | Unifocal | 137 (48) | 43 (15) | 55 (19) | 39 (14) | 0.772 |
|  | Multifocal | 142 (50) | 46 (16) | 55 (19) | 41 (14) |  |
|  | Missing | 4 (1) | 0 (0) | 2 (1) | 2 (1) |  |
| Size <sup>b</sup> | < 3cm | 43 (15) | 16 (6) | 17 (6) | 13 (4) | NA |
|  | ≥ 3cm | 46 (16) | 18 (6) | 18 (6) | 7 (2) |  |
|  | Missing | 194 (69) | 55 (22) | 62 (24) | 13 (22) |  |
| Concomitant CIS | No | 221 (78) | 65 (23) | 93 (33) | 63 (22) | 0.216 |
|  | Yes | 62 (22) | 24 (8) | 19 (7) | 19 (7) |  |
| LVI <sup>c</sup> | No | 239 (84) | 76 (27) | 90 (32) | 73 (26) | 0.417 |
|  | Yes | 11 (4) | 4 (1) | 4 (1) | 3 (1) |  |
|  | NA | 33 (12) | 9 (3) | 18 (6) | 6 (2) |  |
| Variant histology | No | 237 (83) | 71 (25) | 104 (37) | 62 (22) | 0.002 |
|  | Yes | 46 (16) | 18 (6) | 8 (3) | 20 (7) |  |
| EAU Risk group | High-risk | 124 (44) | 37 (13) | 56 (19) | 31 (11) | 0.213 |
|  | Highest-risk | 159 (56) | 52 (18) | 56 (19) | 51 (18) |  |
| BCG failure <sup>d</sup> | No | 156 (55) | 59 (21) | 68 (24) | 29 (10) | <0.001 |
|  | Yes | 127 (45) | 30 (11) | 44 (15) | 53 (19) |  |
| HG-RFS | 1 year (CI) | 73 (67-78) | 77 (68-86) | 76 (68-84) | 64 (55-76) | <0.001 |
| Progression | No | 187 (34) | 68 (24) | 80 (28) | 39 (14) | <0.001 |
|  | Yes | 96 (66) | 21 (7) | 32 (11) | 43 (15) |  |
| PFS | 2 year (CI) | 80 (75-84) | 88 (82-96) | 83 (76-90) | 64 (54-76) | <0.001 |
|  | 5 year (CI) | 70 (65-76) | 76 (64-86) | 76 (69-85) | 55 (46-67) |  |
| Follow-up (months) | Median (IQR) | 73 (51-104) | 76 (56-103) | 75 (55-104) | 68 (29-100) | 0.192 |

BCG = Bacillus Calmette-Guérin; BRS = BCG Response Subtype; CI = confidence interval; CIS = carcinoma in situ; EAU = European Association of Urology; HG-RFS = high-grade recurrence-free survival; IQR = Interquartile range; NA = not applicable; re-TURBT = repeat transurethral resection of bladder tumor before BCG-induction. <sup>a</sup> Percentages may not add up to 100% due to rounding; <sup>b</sup> Not applicable due to missing variables; <sup>c</sup> Assessed only in T1 disease; <sup>d</sup> BCG failure as specified by major urology guidelines, which includes patients with tumors who developed muscle-invasive recurrences, persistent T1HG after BCG-induction and HG recurrences after adequate BCG therapy.

### BRS3 is an aggressive molecular subtype of HR-NMIBC

The BRS clearly delineated a group of patients with a high risk of progression (BRS3). Unraveling the underlying molecular characteristics that drive BRS3 could explain differences in clinical outcome and possibly lead to identification of candidate druggable targets for novel treatments. Based on the overlapping gene expression patterns and PFS, we combined Cohort A and B (*n*=283). Differential gene expression (DGE) analysis showed that BRS3 tumors had overexpression of basal and EMT activation markers (*CD44, KRT5/6, DDR2, VIM, SNAI1, ZEB1/2* and *TWIST1)*. Furthermore, BRS3 tumors were characterized by overexpression of immune suppressive genes, including immune-checkpoint genes (*PD-1/PD-L1* and *CTLA-4*), T regulatory markers (T regs; *CD4/CD25/FOXP3*), myeloid derived suppressor cell (MDSC) markers, colony stimulating factors *(CSFs)* and chemokines *(CXCLs* and *CXCRs)*, which have been implicated in BCG treatment failure [23]. A full list of differentially expressed genes is reported in **Table S3**.

We then analyzed pathways associated with distinct subtypes. Individual genes upon which the BRS pathway activity is based are listed in **Fig. S6A**. In BRS3 tumors, gene set enrichment analysis identified pathways known to be associated with cancer progression, such as EMT, Notch, MAPK and angiogenesis (**Fig. 2A/B**). Moreover, immune-related pathways were strongly enriched, e.g., complement, IL6-JAK-STAT3 and IL2-STAT5 (**Fig. 2A/B**). Complete results are listed in **Table S4**.

**Fig. 2.**
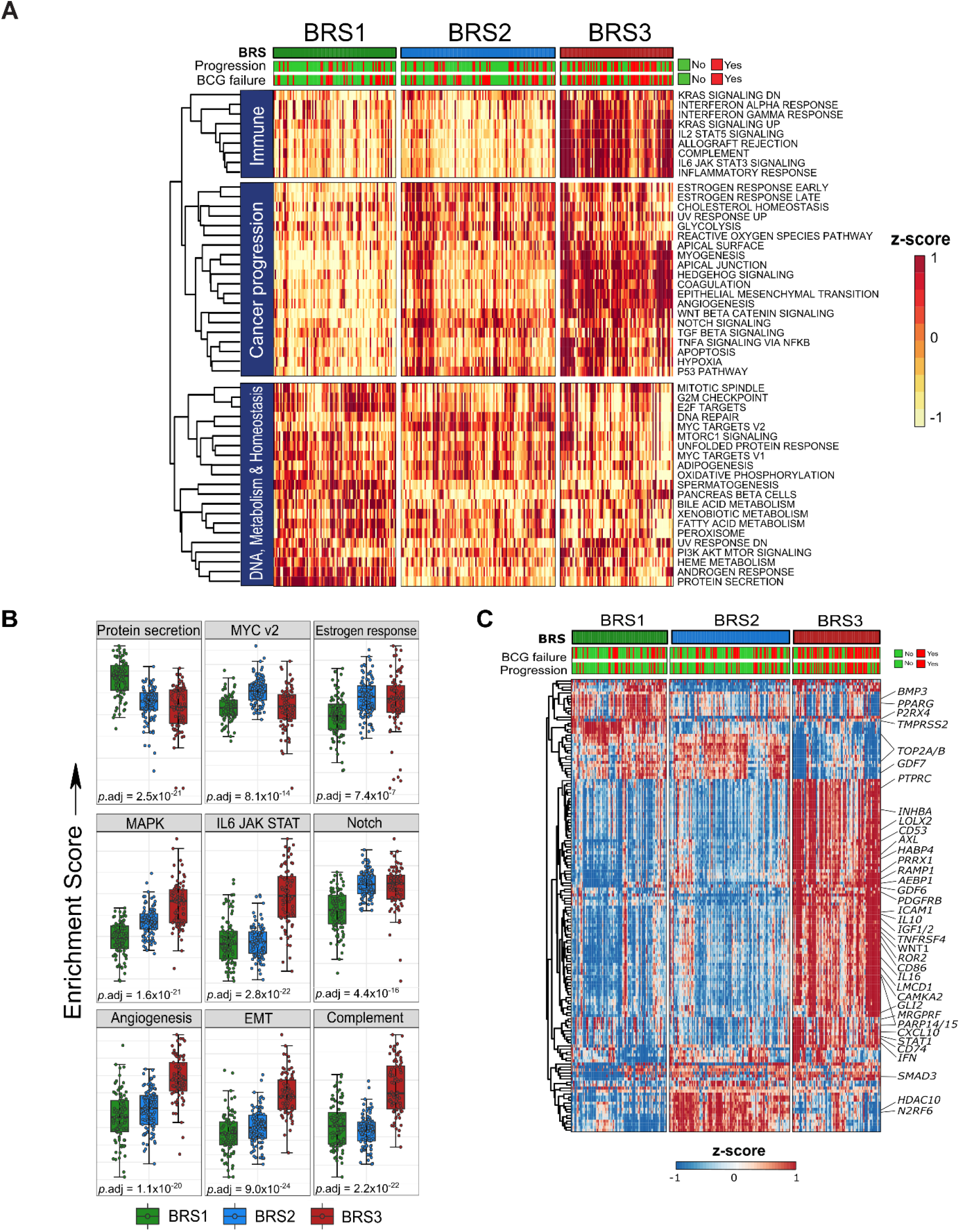
Gene set enrichment analysis and regulon analysis grouped according to the BRS in Cohort A+B. **A:** Heatmap of all 50 GSEA hallmarks in *n*=283 pre-BCG, HR-NMIBC patients. Clustering on gene signatures (rows) indicate the existence of different molecular subtypes in HR-NMIBC. **B:** Boxplots of selected gene signatures in *n*=283 pre-BCG HR-NMIBC patients grouped according to BRS; *p*-values are Kruskall-Wallis tests. **C:** Heatmap of the top 200 most varying regulons using single-sample VIPER analysis on *n*=283 pre-BCG tumors grouped by BRS (columns). Hierarchical clustering between samples confirms three distinct molecular subtypes; key regulators for which small molecule inhibitors exist are highlighted. **Abbreviations**: BCG = Bacillus Calmette-Guérin; BRS = BCG Response subtypes; GSEA = gene set enrichment analysis; HR-NMIBC = high-risk non-muscle invasive bladder cancer; VIPER = Virtual Inference of Protein Activity by Enriched Regulon Analysis.

Regulon analysis using Viper was performed to allow for computational inference of protein activity based on downstream target gene expression patterns in each subtype [24]. Regulons comprising the top 150 most varying transcriptional regulators for which small molecular inhibitors are available are shown for the three HR-NMIBC subtypes (**Fig. 2C**) [25]. These genes may prove to be attractive candidates for subtype-specific drug targeting. Several regulatory genes have previously been identified as important genes driving bladder cancer (BC). These genes have been grouped according to BRS in **Fig. S6B** [18]. We observed low *RB1* and *TP63* in BRS3 tumors, which is often seen in aggressive BC [26]. We confirmed activity of EMT-associated regulons (*ZEB2, TWIST2* and *SNAI3)*, and enrichment of regulators related to MDSCs and B cells (*CD14, CD86*), T regs (*BATF3, IL10, IL16, FOXP3)* and T cell polarization *(STAT1/STAT4* and *TBX21*) (**Table S4**). Taken together, BRS3 tumors were enriched for regulators associated with EMT activation, a basal phenotype and immune checkpoint proteins, and these results associate biological features of BRS3 tumors with the corresponding poor clinical outcome.

### BRS1 and BRS2 HR-NMIBC have luminal characteristics

As with BRS3, we set out to describe relevant molecular features associated with BRS1-2. DGE analysis showed that BRS1-2 tumors had overexpression of the luminal marker *PPARG* (**Fig. S6A, Table S3**). A typical luminal marker is *FGFR3*, but *FGFR3* expression was high in BRS2 tumors only. Pathway analysis revealed BRS1 tumors had upregulated cell cycle and metabolic processes involved in mycobacterial (BCG) processing, such as enrichment of autophagy, ubiquitination and proteasome degradation and protein secretion (**Fig. 2B, Table S4**). These findings provide mechanistic insight into the more favorable outcome associated with BRS1 patients after BCG treatment. GSEA indicated that BRS2 was the least enriched for immune related pathways (**Fig. 2A**) [27]. MYC pathway activity (**Fig. 2B**), the absence of a *carcinoma in situ* (CIS) profile (**Fig. 1C/S3**), enriched luminal signatures (**Fig. S6C**) and a high proportion of pure urothelial cell carcinoma (**Table 1**) were associated with BRS2. Hence, BRS2 tumors align with features seen in papillary BC. Subtype-specific and potential druggable regulatory proteins are listed in **Fig. 2C**. The activity of luminal regulators, *FOXM1, FOXA1, GATA3* and *ERBB3* was high in both BRS1 and BRS2 tumors, but not in BRS3 tumors. Consistent with luminal papillary MIBC, BRS2 tumors retained *SHH* activity (**Fig. S6B**) [18]. Taken together, characterization of BRS1 and BRS2 tumors shows these are molecularly distinct, but both share luminal features, which may explain the better outcomes after BCG.

### The tumor microenvironment of BRS3 displays immune suppressive features

BCG immunotherapy is dependent on interactions with the tumor microenvironment (TME). As findings pointed towards high immunological activity in BRS3 with a poor outcome after BCG treatment, we analysed the TME using transcriptome deconvolution and spatial proteomics. First, we assessed the overall immune infiltration using the ESTIMATE algorithm and confirmed that BRS3 tumors had the highest immune and lowest tumor purity scores as compared to BRS1-2 tumors (**Fig. 3A**) [28]. To gain further insight into immune cell composition of BRSs, we applied the CIBERSORT immune deconvolution method, which allowed for between tumor and between immune cell population comparisons [29]. BRS3 showed the highest infiltration of B cells, tumor-associated macrophages (TAMs) M1/M2, CD8+ T cells and T regs (**Fig. 3B**). Findings were verified using two additional independent algorithms (**Table S5**) [30, 31].

**Fig. 3.**
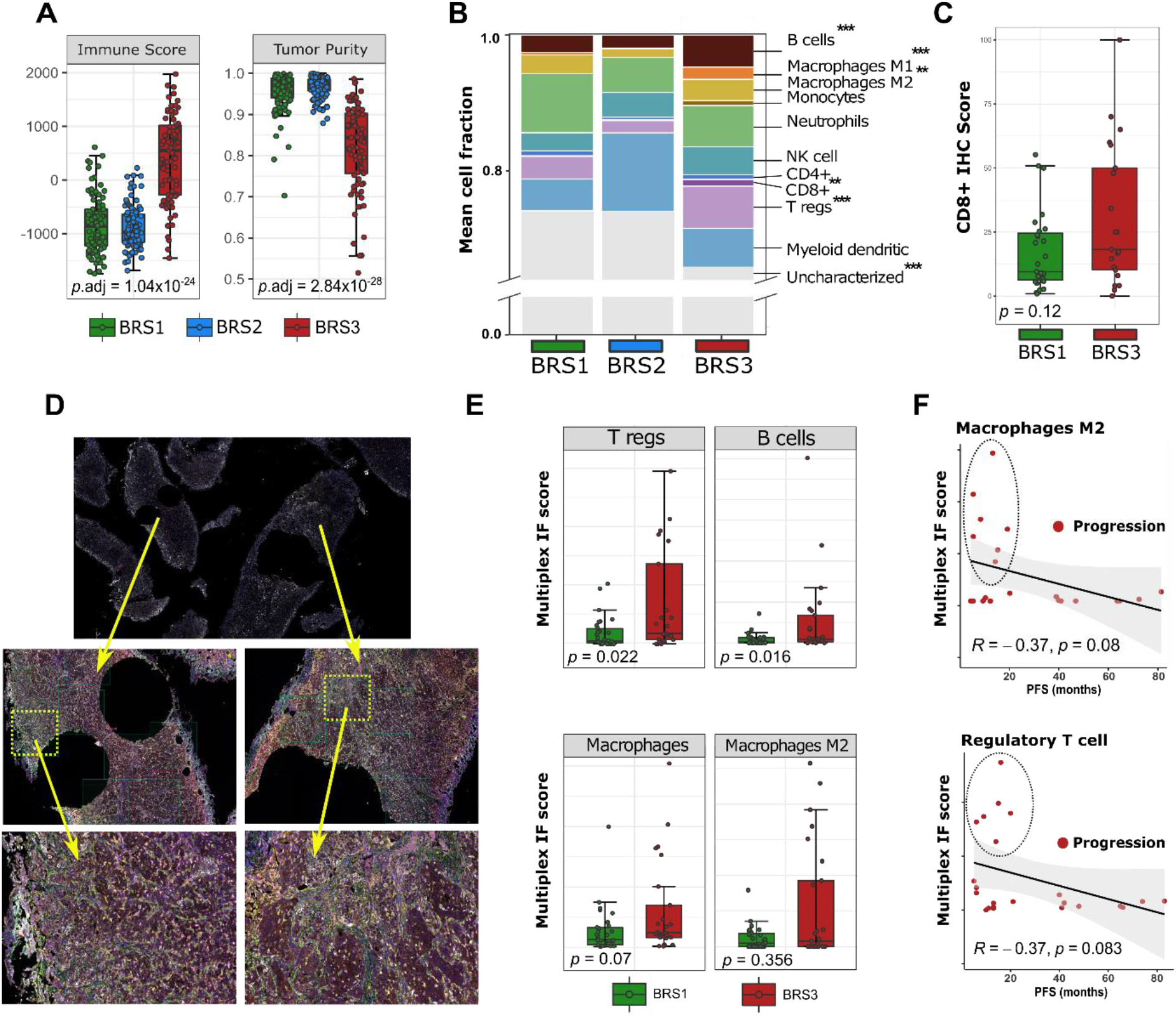
Immune deconvolution of pre-BCG tumors from Cohort A+B and spatial proteomics of the tumor microenvironment grouped by BRS from Cohort A. **A:** Boxplots predicting tumor purity and immune infiltration in *n*=283 pre-BCG tumors grouped by BRS [28]; *p*-values are Kruskal-Wallis tests. **B:** Immune cell deconvolution from RNA-seq: a single column is the sum of ten immune cell subpopulations and non-characterized cells grouped by BRS; *p*-values are Wilcoxon tests for BRS1/2 vs BRS3; \*\**p*.adj<10^−3^; \*\*\**p*.adj<10^−4^. **C:** Immunohistochemistry of CD8+ cytotoxic T cell infiltration in *n*=25 BRS1 and n=27 BRS3 tumors. BSR2 tumors were excluded, as data indicated very low immunological activity in these tumors. **D:** Selection of regions for spatial proteomics; on average 20 regions of pre-BCG BRS1/3 tumors surrounding the macrodissected areas used for RNA-sequencing were selected. Mean intratumoral protein expression was used for analyses. **E:** Boxplots of multiplex immunofluorescence (regulatory T cells, B cells, Macrophages and Macrophages M2) for *n*=27 BRS1 and n=25 BRS3 tumors. **F:** Scatterplot for immunofluorescence results for only patients with progression. Only few patients had tumors that were high in regulatory T cells and macrophages M2, yet all of these patients had early progressive disease (≤2 year). **Abbreviations**: BCG = Bacillus Calmette-Guérin; BRS = BCG response subtypes.

Previous *in vivo* studies demonstrated a critical role for CD8+ T cells in BCG-mediated anti-tumor immunity [32, 33]. Therefore, high infiltration of CD8+ T cells in treatment-naive BRS3 tumors seemed a paradoxical finding, as most patients with BRS3 did not respond to BCG treatment. Thus, we investigated if BRS3 tumors have high CD8+ T cells at the protein level. In Cohort A, all BCG responders that were BRS1 (n=27) were selected, and from all non-responders, we selected BRS3 tumors (n=25). The luminal *FGFR3* expressing BRS2 subtype was excluded, as our expression data indicated that BRS2 tumors had the lowest expression of immune-related genes. Immunohistochemistry was successfully performed on 48 selected pre-BCG BRS1/3 tumors with a clinically approved antibody for CD8+ T cells. Consistent with RNA expression data, we detected more intratumoral CD8+ T cells in BRS3 vs. BRS1 tumors (*p*=0.12, **Fig. 3C**), but results were not statistically significant. The same tumors were also investigated by multiplex proteomics for CD4+, T regs, B cells and Macrophages (M1/M2) and Vimentin (VIM). Areas surrounding the macrodissected tumor punches used for RNA isolation were selected (**Fig. 3D**). We confirmed an increased number of tumor infiltrating immune cells (CD4+ [*p*=0.021], T regs, B cells and Macrophages) and a higher expression of intratumoral VIM (*p*<0.001) in BRS3 on a protein level as compared to BRS1 using automated quantification on tumoral vs stromal tissue (**Fig. 3E**). Interestingly, in several patients with early progressive disease, high infiltration with Macrophages M2 and T regs was seen, suggesting that this might serve as marker for treatment failure in a subset of patients (**Fig. 3F**). Taken together, despite a clear trend towards higher CD8+ T cells and Macrophages on RNA-level, results were not significantly recapitulated at the protein level, possibly due to sample size. Nonetheless, BRS3 tumors were more infiltrated with T regs and B cells, both associated with immune suppression, and these findings have previously been linked to a poor clinical outcome after BCG [34, 35].

### BRS improves current clinical risk stratification of HR-NMIBC

We investigated how the BRSs impact BCG response and clinical outcome in Cohort A+B. According to international recommendations, BCG response is defined as being free of high-grade (HG) disease after 6 months of adequate BCG exposure [36]. Using this endpoint, a differential response to BCG treatment was observed between subtypes (BRS1 85%, BRS2 82%, BRS3 68%; *p*=0.017). In addition to PFS (**Fig. 4A**), patients with BRS3 tumors had a poorer HG recurrence-free survival (**Fig. 4B**). In multivariate analyses, BRS3 vs. BRS1/2 was an independent predictor of PFS (HR 2.7, *p*<0.001, **Fig. 4C**).

**Fig. 4.**
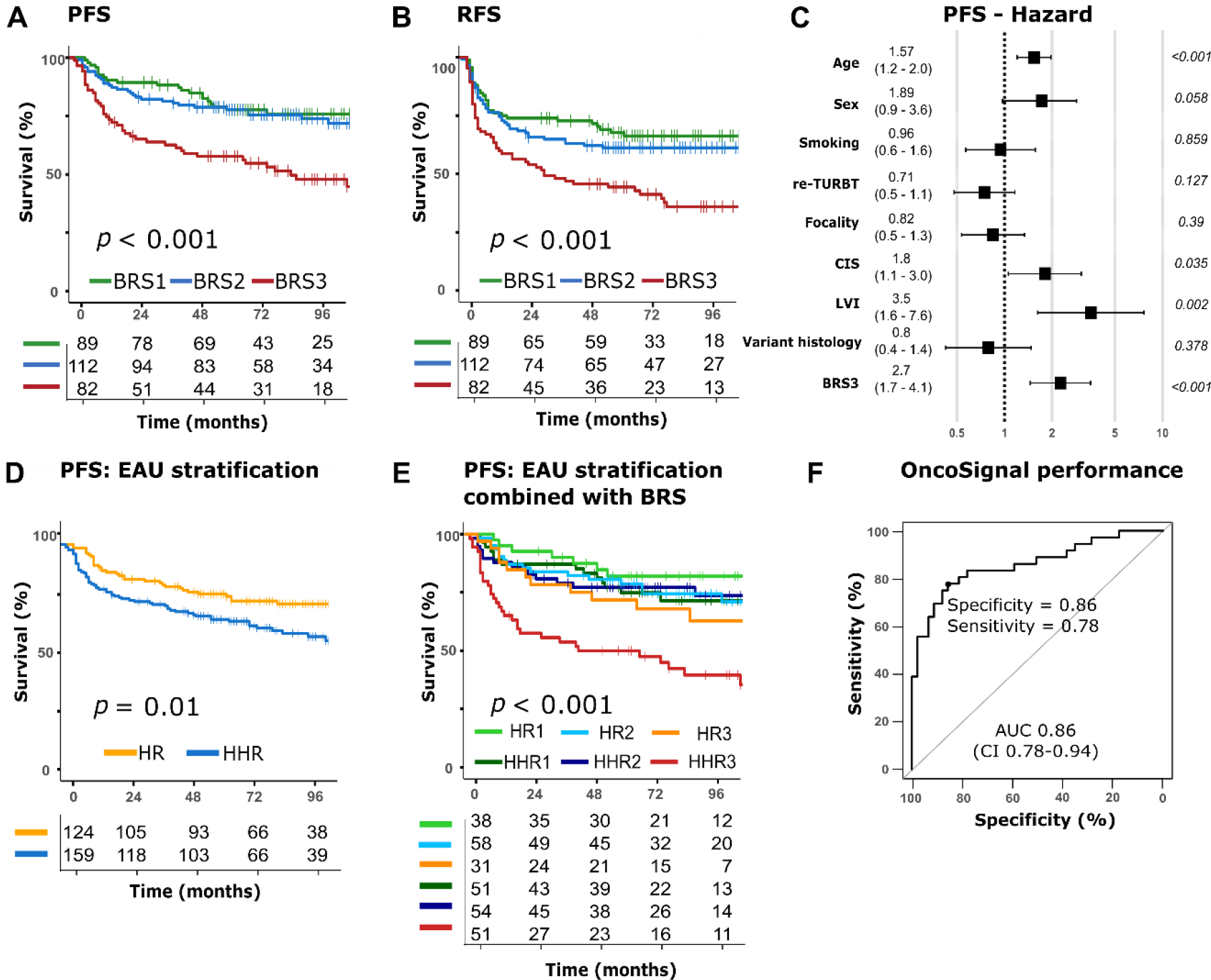
Kaplan-Meier estimates of survival according to the BRS in *n*=283 pre-BCG HR-NMIBC patients (Cohort A+B) and assay performance to predict BRS3 in Cohort A. **A:** Progression-free survival (PFS) stratified according to BRS. **B:** Recurrence-free survival (RFS) for high-grade tumor recurrences stratified according to BRS. **C:** Forest plot of multivariable Cox regression analysis; BRS3 vs BRS1 + BRS2. **D:** PFS stratified for the current EAU guideline recommendations (high-risk [HR] patients vs subgroup of very high-risk [HHR] patients). **E:** PFS stratified for current EAU risk-stratification and the BRSs. **F:** Subtype prediction for BRS3 vs BRS1/2 based on the assay results in Cohort A. Figure depicts the ROC with the area under the curve based on a logistic regression model, with corresponding test sensitivity and specificity reported at a threshold of 0.385. **Abbreviations**: BCG = Bacillus Calmette-Guérin; BRS = BCG response subtypes; EAU = European Association of Urology; HR-NMIBC = high-risk non-muscle invasive bladder cancer; Progression free survival (PFS).

Next, we analyzed the added value of the BRS classifier to the clinicopathological risk stratification of HR-NMIBC recommended by the EAU guidelines [1]. This risk stratification is based on the presence of stage T1, grade 3 or high-grade, concurrent CIS, multifocality, large tumors (≥3cm), aggressive forms of variant histology and lymphovascular invasion which are associated with a high risk (HR) or a very high risk of progression (HHR) [1]. As HHR patients have a higher risk of progression than HR patients, guidelines recommend consideration of an early RC (instead of BCG). In our study, HHR patients had a 2-year PFS of 75% vs 85% for HR patients (*p*=0.01, **Fig. 4D**). Combining the BRS with the EAU HR-NMIBC risk model improved risk stratification by identification of a subgroup of patients with the highest risk of progression (HHR3) (**Fig. 4E**). HHR3 patients had a 2-year PFS of only 54%. Importantly, HHR1 vs HHR2 patients, normally also considered for early RC, now have a similar 2-year PFS as HR patients. Considering the strong findings on BRS combined with clinicopathological features to predict outcome, we developed a preliminary nomogram for the research community to estimate the risk of progression, which should be validated prospectively (**Fig. S7A**). Finally, we assessed whether previously reported RNA-subtypes were able to predict response to BCG treatment in our total Cohort A+B. None of the published MIBC or NMIBC subtypes could identify a clinically relevant subset of patients with the highest risk of progression (**Fig. S8A-G)**.

### BRS3 patients can be identified with a commercially available test

To investigate the clinical utility of the BRS, we assessed whether a commercially approved assay (OncoSignal®) was able to identify BRS3 versus BRS1-2 tumors. The assay is qPCR-based and measures seven signal transduction pathways (details in methods) [37]. The same RNA used for transcriptomic sequencing of Cohort A was selected for OncoSignal® analysis. The assay was able to distinguish BRS3 from BRS1/2 tumors with BRS3 having high signal transduction activity of MAPK, ER, and Notch pathways (**Fig. S7B**). Slight differences between BRS3 and BRS1-2 were found based on Hedgehog, PI3K and TGFβ pathways. From a clinical perspective, it is important to distinguish aggressive BRS3 tumors from both BRS1/2 tumors, as BRS3 tumors might benefit from a different therapeutic strategy than standard-of-care. Thus, based on the pathway activity scores, we developed a logistic regression model to predict if a tumor was BRS3. The model resulted in strong overall performance (AUC 0.86), with optimized thresholds resulting in a true positive rate of 78% and true negative rate of 86% (**Fig. 4F**). Although findings need validation in an independent cohort, we have established for the first time that a commercially available assay can be used to identify a clinically relevant aggressive type of HR-NMIBC.

### Post-BCG tumor recurrences are enriched for BRS3

To gain further insight into molecular changes that occur after BCG treatment failure, we applied the BRS classifier to 44 post-BCG tumors (34 pre/post-treatment matched tumors). Strikingly, BRS3 predicted tumors were enriched in post-BCG 28/44 (64%) recurrences vs pre-BCG tumors 82/283 (28%) (**Fig. 5A**). From the 34 matched tumors, 26/34 patients had BRS1-*2* tumors pre-BCG of which 14/26 tumors switched to BRS3 post-BCG.

**Fig. 5.**
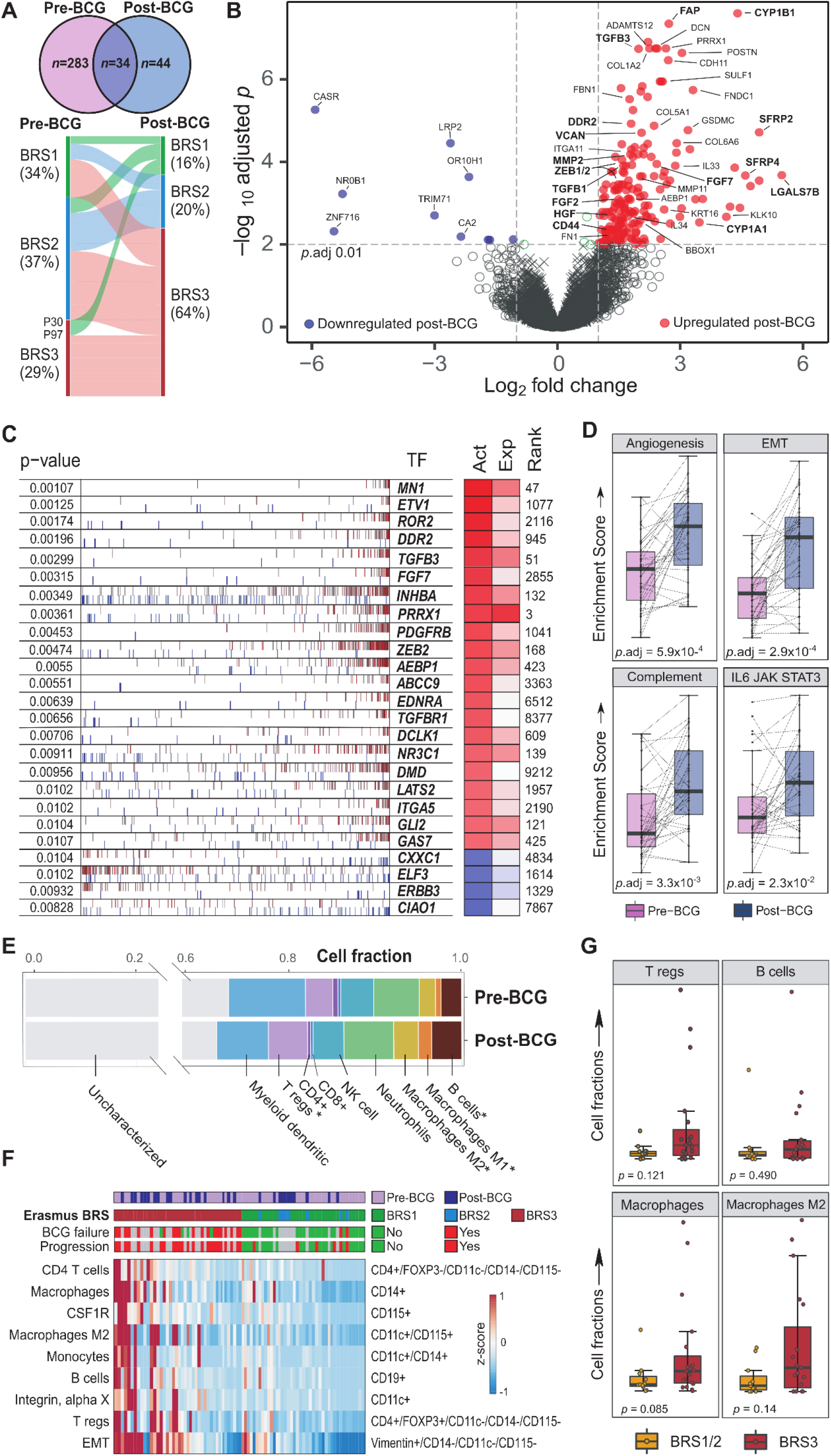
Pre- vs post-BCG transcriptomic and spatial proteomic comparisons. **A:** Frequency of BRS in pre-BGC vs post-BCG tumors in Cohort A+B. Alluvial plot details 34 patients with matched pre- and post-BCG tumor samples. **B:** Volcano plot of differentially expressed mRNAs in matched post-BCG and pre-BCG tumors from *n*=34 patients. **C:** Top regulons with small molecule inhibitors that are up- or down-regulated in pre-BCG tumors and post-BCG tumors depicted. In red: regulators enriched in post-BCG recurrences. In blue: regulators enriched in pre-BCG tumors. **D:** Selected gene set enrichment hallmarks comparing *n*=34 paired pre- and post-BCG tumors samples. Dashed lines represent pathway activity scores between paired samples of a single patient; *p*-values are paired Wilcoxon tests. **E:** Immune cell deconvolution based on RNA-seq: a single column is the sum of ten different immune cell populations and non-characterized cells found in the tumor microenvironment grouped by pre-BCG tumors or post-BCG recurrences; *p*-values are Wilxocon tests; \**p*.adj<0.05. **F:** Heatmap with spatial proteomics grouped by subtype and sorted by z-score. Results indicate an increased presence of tumor infiltrating immune cells in BRS3 vs BRS1/2 tumors and post-BCG vs pre-BCG tumors. **G:** Proteomic results of *n*=30 post-BCG recurrences grouped by post-BCG BRS. BRS3 tumors had a non-significant increased number regulatory T cells and Macrophages (M2). **Abbreviations**: BCG = Bacillus Calmette-Guérin; BRS = BCG response subtypes.

Overexpressed genes in post-BCG samples can indicate mechanisms involved in BCG treatment failure and might therefore provide candidate target genes for development of novel treatments. Paired DGE analysis revealed that in post-BCG tumors, *CYP1A1* and *CYP1B1* were highly overexpressed, both key genes involved in phase-1 drug metabolism (modification), as well as genes essential for attachment of BCG to urothelial cells (*FN1, FNDC1, ITGA5*). These results in tumors that do not respond to treatment, indicate that overexpression could be a consequence of ineffective mycobacterial antigen exposure to urothelium. EMT related genes (*ZEBs, TGFβ1/3*), growth factors (*FGF2, FGF7, HGF*) and genes associated with basal-like disease and cancer progression (*CD44, KRT6/16, LGALS7)* were overexpressed (**Fig. 5B**) [38-40]. Collagen receptor *DDR2* and EMC remodeling genes (*MMPs, ADAMTSs, VCAN, FAP*) were also highly overexpressed after treatment. Although these genes are involved in wound healing and tumor scarring, extensive preclinical work has shown that they promote bladder cancer metastases and are predictive of a poor outcome [41-43]. Additionally, post-BCG tumors showed increased expression of *SFRPs –* WNT antagonists *–* which is consistent with the WNT pathway activity found in pre-BCG BRS3 tumors and might indicate WNT signaling involvement in BCG resistant tumors [44]. Overall, of the multiple genes differentially overexpressed in tumors displaying the pre-BCG BRS3 gene signature, most were overexpressed in tumors that failed treatment and described in the literature to be associated with cancer progression (**Table S6**). Many of these genes can be targeted by available small molecules, providing an excellent starting point for development of novel treatments in these BCG-resistant patients [25].

Next, we investigated consistently expressed gene fusions, important regulatory networks, pathways and non-synonymous SNVs to investigate which potentially actionable mechanisms underlie recurring disease post-BCG. We identified gene fusions that were present before and after treatment within the same patient. Although infrequent, druggable gene fusions such as *FGFR3--TACC3* fusions were detected in three patients (**Fig. S9A, Table S7**). Transcriptional regulators that were enriched in post-BCG tumors include *DDR2, TGFβs, AEBP1*, glucocorticoid receptor *NR3C1* and Fibroblast growth factor *FGF7* (**Fig. 5C, Table S6**). By performing a within-patient pathway analysis, we noted enrichment of EMT, complement, IL6-JAK-STAT3 and angiogenesis in post-BCG recurring tumors (**Fig. 5D; Table S6**), supporting the hypothesis that recurrences have a more aggressive biology. Signatures that contributed to the pre-BCG BRS3 were also the dominant signatures present in recurring tumors (**Fig. S9B, Table S6**). Variant calling identified significant mutational differences between pre- and post-BCG tumors. Interestingly, a dozen of these genes belonged to EMC organization, which is interesting as ECM signatures were overexpressed in treatment failures as well (**Fig. S9C; Table S6**). To summarize, post-BCG tumors displayed enrichment of pathways that are involved in cancer progression. Additionally, several promising actionable targets in post-BCG recurrences were detected.

Finally, we investigated the TME of post-BCG recurrences. Immune deconvolution on matched samples showed more T regs, Macrophages and B cells in post-BCG tumors (**Fig. 5E**). Intratumoral multiplex spatial proteomics of *n*=85 samples showed an increased presence of immune subpopulations in post-BCG BRS3 **(Fig. 5F**). From the post-BCG recurrences, only BRS3 tumors showed a trend towards increased numbers of regulatory T cells and Macrophages (M2) as compared to BRS1-2 tumors (**Fig. 5G**). Protein analysis pointed towards a phenotype that is associated with immune suppression in tumors that fail BCG treatment.

Altogether, these data strongly support the hypothesis that BRS3 tumors have a clinically and biologically aggressive phenotype. In addition, paired analyses of tumors that did not respond to BCG provide targetable gene candidates for preclinical research into alternative treatments for BCG non-responsive tumors.

## Discussion

We analyzed the transcriptomes of HR-NMIBC patients to characterize molecular features associated with response to BCG. This is the largest study to date in BCG-treated HR-NMIBC patients that establishes the existence of molecular subtypes predictive of outcome after BCG. The robustness of the BRS classifier was validated in a second profiled HR-NMIBC patient cohort. We identify BRS3 as a clinically relevant subgroup of patients, who have enrichment of EMT activation markers, a basal and immunosuppressive phenotype, overexpression of immune checkpoint genes and a poor progression-free survival as compared to BRS1 and BRS2 patients. Addition of BRS to guideline-based patient risk groups improved stratification of patients with the highest risk of progressive disease. Current guideline recommendation on disease management of HR-NMIBC patients is to consider an early RC for tumors with very-high risk clinicopathological criteria. Molecular subtyping in very-high risk BRS1 and BRS2 patients have a significantly lower risk of progressive disease, which creates the possibility for these patients to preserve their bladder. In contrast, the addition of BRS3 to the very high-risk group showed an even poorer PFS, which suggests that these patients should strongly be considered for early RC.

BCG treatment failure in 30-50% of patients is a major risk factor for progressive disease and death from BC. Thus, much effort has been made to identify patients that do not benefit from treatment and to search for treatment alternatives to radical surgery. Previous RNA-based molecular subtyping studies were not associated with clinical outcome after BCG treatment, possibly because these studies lacked sufficient numbers of patients or were difficult to interpret in the clinic due to the high number of subtypes [14]. UROMOL21 did identify three T1HG subtypes, but these subtypes were not able to predict failure of BCG treatment [13]. The T1BC subtyper was built for recurring disease and developed in a population with a limited number of patients who developed progression [17].

Previous work showed that the presence of tumor associated macrophages and *FOXP3*-positive T regs in the TME are associated with worse outcome in BCG-treated BC [45]. Kates *et al*. found that BC progression occurred in BCG treated immunocompetent rats with an increased presence of *FOXP3*-positive T regs and rats having a decreased adaptive immunity [34]. BRS3 tumors also showed signs of CD8+ T cell infiltration, Th1 polarization and interferon-γ pathway activity, vital for anti-tumor immunity and BCG effectiveness [46, 47]. This is a seemingly paradoxical finding as BRS3 tumors have the worst clinical outcome, but strong Th1 predisposition should be counterbalanced to avoid auto-immunity, which may explain the T regs, B cells and Macrophages in BRS3 tumors [48]. The IL2-STAT5 pathway was enriched in BRS3 tumors. Measurement of urinary IL-2 during and after BCG treatment has been proposed as a marker of response to BCG [49, 50]. However, IL-2 also plays an important role in T reg development via *STAT5* [51]. Because of the high CD8+ T cell activity in BRS3 tumors and the non-response to BCG immunotherapy, results might be suggestive of T cell exhaustion in BRS3 tumors. Previous work indicates that T cell exhaustion may lead to an ineffective cascade of additional anti-tumor immunity, which might play a role in diminished therapeutic success of BCG in BRS3 tumors [52, 53].

A strong aspect of this study is the availability of matching pre- and post-BCG treatment samples, which allows for the unique investigation of overexpressed genes in BCG treatment failures. For instance, previous work showed that TGF-β attenuates anti-PD-L1 therapy outcomes by exclusion of cytotoxic T cells from the TME [54]. Here, high expression of TGF-β was demonstrated in post-BCG tumors (mostly BRS3), suggesting that TGF-β is a key player to overcome BCG-resistance and should be investigated as a druggable target. Overexpression of checkpoint genes in BRS3 tumors (PD-(L)1, CTLA-4, etc.) in our study provides a rationale for the use of immune checkpoint inhibition in BCG-unresponsive patients, as observed in the Keynote-057 clinical trial where patients had a durable response to anti-PD-1 [55]. Recently, anti-PD-1 combined with anti-CTLA-4 showed excellent response rates as neoadjuvant therapy in MIBC, and the data indicates this strategy has potential in BCG-unresponsive disease as well [56]. Lastly, *in vivo* studies showed positive synergistic effects on tumor load and increased CD8+ T cell influx when dasatinib (anti-DDR2) was combined PD-L1 blockade [57]. In addition to RNA overexpression of checkpoint proteins in BRS3, we demonstrate an increased expression of *DDR2* in post-BCG tumors, thus supporting the potential of anti-PD-L1 combination treatment with anti-DDR2. The latter mechanism is in part due to M2 macrophages via CCL2, in which CCL2 inhibition could improve checkpoint therapy [57, 58].

BRS1 tumors had the most favorable PFS and showed molecular similarities to the genomic subtype, GS2, previously identified by Hurst *et al*. in stage Ta/low-grade BC [12]. BRS1 tumors had increased expression of genes related to intracellular protein trafficking and antigen presentation, which is vital for mycobacterial processing, and MHC1-mediated cytotoxic T cell killing of cancer cells [59-63]. Enhanced autophagy has been found to be important in processing of BCG and improved antigen presentation *in vivo* [64, 65]. SNPs in *ATG5* and *ATG2B* negatively influence trained immunity after restimulation with BCG and inhibition of autophagy directly attenuated bacterial processing and antigen presentation [66]. These results led us to hypothesize that enhanced autophagy during BCG treatment will improve the immune response.

BRS2 tumors were luminal, with high *FGFR3* expression, activation of MYC, but substantial within-subtype heterogeneity. Luminal heterogeneity is a consistent finding in BC and complicates clinical implementation of subtyping [67]. The relationship of *MYC* to *FGFR3* activity was elucidated *in vitro*, in which Mahe *et al*. discovered that *FGFR3* expression led to downstream MYC accumulation and increased *FGFR3* activity in a positive feedback loop [68]. Recently, anti-FGFR treatment in metastatic BC was shown effective with low toxicity rates [69]. Therapeutic inhibition of FGFR is an option that has potential therapeutic translation in BRS2 tumors given the high prevalence of FGFR alterations in NMIBC and high *FGFR3* activity in BRS2 tumors. Of interest was *N2RF6* regulon activity in BRS2 tumors. Recently, it was shown that loss of *N2RF6* in a CRISPR/Cas9 mouse model leads to hyper-responsive T cells and enhanced responses to PD-L1 inhibition [70, 71]. It would stand to reason that blocking *N2RF6* could sensitize BRS2 tumors to immune-checkpoint inhibition.

Although next-generation sequencing data is more easily accessible, molecular subtyping based on transcriptomics remains time-consuming, expensive and requires dedicated personnel. To pave the way for clinical translation of our findings, we used a commercially approved qPCR-based assay and show that this test is able to accurately identify BRS3 HR-NMIBC patients. As a next step, a non-randomized phase 2 study is needed to assess the predictive value of BSR3 in a prospective manner. To this end, a comparison between the BRS transcriptomic classifier and the commercial assay is required to test whether OncoSignal can be widely used as a means to stratify HR-NMIBC patients. Alternatives to BCG treatment based on assay results can also be investigated. Clinical trials targeting signal transduction pathways, such as MAPK and Notch are ongoing, and based on our findings, these pathways are candidates for further investigation in BRS3 tumors [72, 73]. Finally, it would be interesting to enhance assay stratification by inclusion of immunological pathways such as JAK-STAT, which was highly upregulated in (post-BCG) BRS3 patients and is also considered as target for pathway-directed precision medicine [74].

This study is not without limitations. First, tumors from BCG non-responders were overrepresented as compared to a real-world situation [75]. For the discovery cohort, we specifically selected true BCG responders and BCG non-responders, because the objective was to find molecular characteristics associated with response to BCG. Second, the OncoSignal assay was run on the same RNA as we performed RNA-sequencing on. While comparison between these assays is a necessary first step, independent validation using RNA from an independent cohort must be done. Third, future studies should address the issue of RNA-based spatial heterogeneity, as false positive and false negative subtypes might be a result of sample collection bias or inherent BC subtype heterogeneity [76, 77]. Assessment of histological and molecular heterogeneity will likely help to explain BCG-resistance due to selection pressure, which may have prognostic and therapeutic implications [78, 79]. We found that the proportion of BSR3 increased in post-BCG recurrences. Therefore, we speculate that progression from BRS1/2 to BRS3 after treatment failure is due to tumor plasticity and clonal expansion of aggressive tumor cells.

To conclude, our findings showed the prognostic relevance of molecular subtypes in HR-NMIBC patients. We showed that subtyping can improve the clinical risk stratification and we translate our findings via OncoSignal®, a consumer-ready pathway activity diagnostic test. In addition, the data presented provided unique pre- and post-treatment evidence for development of novel targeted therapies, targeting tumors that fail BCG treatment. Identification of BRS3 tumors may be a critical step for implementation of more aggressive therapeutic regimens such as an early RC or recruitment into clinical trials.

## Materials and Methods

### Patients & Pathology

Tumor samples from primary HR-NMIBC patients who had received ≥5/6 BCG-induction instillations between 2000-2018 at four different Dutch hospitals (Erasmus University Medical Center Rotterdam, Franciscus Gasthuis and Vlietland Rotterdam, Amphia Breda and Reinier de Graaf Gasthuis, Delft) and one Norwegian hospital (Stavanger University Hospital) were compiled. IRB approval was obtained from the Erasmus MC Medical Research Committee (MEC-2018-1097). The regimen of BCG instillations was according to the *Southwest Oncology Group* (SWOG) protocol; clinical follow-up was according to the *European Association of Urology* (EAU) guidelines [1]. Formalin fixed paraffin-embedded (FFPE) material, including TURBTs, re-TURBTs, tumor recurrences, random biopsies, radical cystectomies (RC), pelvic lymph node dissections (PLND) and distant metastases were reviewed by an expert uropathologist (R.F.H.) in accordance with WHO standards for classification of the urinary system and was previously published [3, 80]. Patients were classified as high risk or as very high risk subgroup according to the EAU guidelines on NMIBC. Very high risk criteria included: multiple and/or large tumors (≥3cm), and/or concomitant CIS, and/or lymphovascular invasion (LVI) and/or certain aggressive forms of variant histology (e.g. micropapillary, sarcamatoid etc.) [1].

### Definitions & Statistics

Cohort A consisted of *n*=63 BCG responders and *n*=69 BCG non-responders. Response to BCG was defined as the absence of a high-grade recurrence following at least 5/6 BCG induction instillations and ≥9 BCG maintenance instillations (*i*.*e*. 1-year schedule with at least 3 cycles of BCG maintenance). Cohort A had a median of 18 instillations. Non-response to BCG was defined as the development of one of the following: i) biopsy-proven muscle-invasive bladder cancer, ii) persistent T1HG NMIBC after BCG induction, iii) high-grade NMIBC after adequate BCG therapy, which was defined as ≥5/6 BCG induction instillations plus ≥2/3 BCG maintenance instillations. Only patients were included in whom pathology review showed invasion of high-grade (HG) urothelial cancer cells into the lamina propria (T1HG NMIBC) with absence of cancer cells in the detrusor muscle of the primary TURBT or re-TURBT. BCG-naive tumor samples were selected for whole transcriptome analysis from both BCG responders and non-responders, and from the latter group, we additionally included samples that were the first HG tumor recurrences during BCG therapy, which could be TaT1HG or T2. For Cohort B, additional patients with HR-NMIBC were included. Cohort B resembled Cohort A, with the aim to have a similar number of tumors that responded to BCG (n=88) and that did not respond to BCG (n=63). To increase the number patients in Cohort B, we also included patients with Ta tumors. In Cohort B, patients received ≥5/6 BCG induction instillations, with a median of 13 instillations.

The primary end point was the progression-free survival (PFS), defined as the time from HR-NMIBC diagnosis until development of MIBC, lymph node (LN) or distant metastatic (M+) disease. Secondary end point was the high-grade recurrence-free survival (HG-RFS), defined as the time from HR-NMIBC diagnosis until a biopsy-proven HG recurrence had occured. Disease-specific survival (DSS) was defined as time from HR-NMIBC diagnosis until death of BC. Patients who were lost to follow-up were censored at the last date of follow-up or death. For the between subtype comparisons, a Fisher’s Exact Test for categorical data or a Kruskall-Wallis test for non-parametric continuous data was used. Kaplan-Meier (KM) estimation coupled with the log-rank statistical test was used to model survival over time with KM plots truncated at 8 years. A multivariable Cox-proportional hazard model was generated to determine whether BRS was associated with PFS. Model variables included age, sex, re-TURBT, tumor focality, CIS, LVI and BRSs (BRS1 + BRS2 vs BRS3). Tumor size was excluded due to missing variables. To prevent understaging of primary T1 disease and to exclude bias from absence of a re-TURBT, additional analyses of Cohort A was performed on patients only with a re-TURBT available. For all between subtype and survival analyses, results were determined to be significant at *p*<0.05.

### Clustering analysis pipeline and molecular clustering

Details on whole-transcriptome sequencing and tissue selection are in the supplementary materials and methods. The R package *cluster* was used to split (3:1) Cohort A based on two 2 variables (BCG-failure [yes/no] and Progression [yes/no]) [81]. The training cohort (*n*=99) was filtered for protein coding transcripts (R package *AnnotationDbi*) [82]. We also removed all immune related genes (union of the signature genes from quanTIseq, EPIC and CIBERSORT algorithms, see following section) from our dataset [29-31]. This strategy prevents predominant clustering on immune related genes and enables between cluster comparisons using RNA-seq based immune cell deconvolution, which makes use of the filtered signature genes. Then, we performed unsupervised clustering (R package *ConsensusClusterPlus*) with top variant genes (500-6000) with steps of 250 genes ranked based off of the calculated row variance [83]. Arguments for ConsensusClusterPlus included: partitioning around medoids (PAM), Pearson correlations and iterating 2500 times with 95% samples. We determined that clustering with the 2000 top variant mRNAs resulted in the most robust sample allocation for three clusters (**Fig. S1B**) as determined by intercluster variation defined by the CDF plot, consensus matrix, and leading to clear differences in PFS (**Fig. S1C**). After analyzing survival and interpretation of GSEA results between clusters, we deemed three gene clusters as clinically informative and useful. Clustering labels from the training cohort was used to produce a multiclass predictive gene signature with the shrunken nearest centroid method (R package *pamr*) [84]. The resulting pamr object was able to predict three classes in the testing cohort (*n*=33). Based on estimated survival (**Fig. S1D**) and overlapping GSEA hallmarks between identical subtypes from both training and testing cohorts (**Fig. S1E**), we confirmed the discovery of three distinct molecular subtypes in Cohort A.

The next step in establishing the algorithm parameters was clustering of all pre-BCG Cohort A patients using the established input parameters. To this end, Cohort B was quantile normalized and matched to Cohort A (the top 2000 most varying genes between samples). Results of ConsensusClusterPlus were used to investigate three, four and five gene clusters (**Fig. S1F**). We again determined that the 3-cluster solution was again the best fit for the data based on intercluster variation through internal validation, differences in GSEA pathway analyses between clusters and survival analysis. We generated a *pamr*-based nearest-centroid gene classifier based on the PAM model from Cohort A using an optimal threshold of 0.1652. This threshold was chosen as this produced the least misclassifications. We locked our model and set out to validate the BRS in Cohort B.

### Subtype validation and calculating molecular signatures

The BRS classifier was used to predict the BRSs for Cohort B. Similarly, the BRS classifier was also applied to the BCG samples from UROMOL21, to the T1BC dataset (GSE154261) and the TGCA dataset [13]. The TCGA BLCA dataset was downloaded using the Genomic Data Commons (GDC) portal (via https://portal.gdc.cancer.gov/projects/TCGA-BLCA). For both datasets, raw counts were VST normalized using DESeq2 and then used for downstream analyses. Using the consensusMIBC *R* package, we generated all MIBC subtypes from our expression matrices [85]. A 68-gene CIS signature score was also built based on a previously reported study from *Dyrskjot et al*. [86]. The luminal BC signature (**Fig S6C**) included the following genes: *FGFR3, PPARG, FOXA1, GATA3, ERBB2 ERBB3, DDR1, UPK1A, UPK2, UPK3A, UPK3B, KRT18, KRT19, KRT20, CDH1, FABP4, CD24, XBP1* and *CYP2J2*. For T1 classification, the *classifyT1BC* R package was used [17]. Results are depicted with Kaplan-Meier estimates, boxplots and heatmaps. All generated heatmaps were ordered according to the following signatures: 1) BRS, 2) Consensus MIBC, 3) TCGA, 4) Lund and 5) MDA subtypes.

### Code availability

Coefficients of the BRS classifier and datasets are found in the *R* package on GitHub (https://github.com/CostelloLab/BRSpred) and supplied in the supplementary data. Gene expression data matrices can be supplied to the BRS classifier. If missing genes are detected, mean values are imputed.

### Immunohistochemistry

Sections were obtained from the same FFPE blocks as used for RNA isolation. Immunohistochemistry was performed according to the BenchMark ULTRA (Ventana) protocol. Sections were deparaffinated and washed (EZ Prep, Ventan). Heating and incubation was performed according to the Ultra-CC1 condition (Cell Conditioning 1, Ventana, 32 minutes at 100 °C) with Reaction Buffer (10X, Tris-based buffer solution (pH 7.6 +/-0.2) washing steps. Next, the primary diagnostics-approved antibody (anti-CD8, clone SP57, Ventana) was added and incubated (32 minutes at 36 °C). Antibody detection was done with the OptiView DAB IHC Detection Kit according to standard diagnostics protocol. For analyses, two independent researchers selected up to six regions surrounding the macrodissected areas used for RNA-sequencing, and counted the CD8+ positive T cells within a single microscopic high-powered field (40x). Boxplots were generated from the mean number of CD8+ positive T cells of up to twelve regions (six regions times two investigators). Wilcoxon testing was done, with statistical significance set at *p* < 0.05.

### Spatial Proteomics

Multispectral imaging was done using the Akoya Vectra Polaris instrument at the Human Immune Monitoring Shared Resource (HIMSR) at the University of Colorado Anschutz Medical Campus. Tissue sections from the same tumor blocks as used for RNA-isolation were stained consecutively with specific primary antibodies according to standard protocols provided by Akoya and performed routinely by the HIMSR. Briefly, the slides were deparaffinized, heat treated in antigen retrieval buffer, blocked, and incubated with primary antibodies for CD19, CD4, CD11c, FOXP3, CSFR1 (CD115), CD14, Vimentin, and pan-CK followed by horseradish peroxidase (HRP)-conjugated secondary antibody polymer, and HRP-reactive OPAL fluorescent reagents. To prevent further deposition of fluorescent dyes in subsequent staining steps, the slides were stripped in between each stain with heat treatment in antigen retrieval buffer. Whole slide scans were collected using the 10x objective with a 1 micron resolution. Regions of interest surrounding the macrodissected areas used for RNA-sequencing were selected for multispectral imaging and quantification tumor areas (3 – 28 fields per tissue, depending on tissue size). Multispectral imaging was performed using the 20x objective with a 0.5 micron resolution. The 9 color images were analyzed with inForm software version 2.5.1 (Akoya Biosciences) to unmix adjacent fluorochromes, subtract autofluorescence, segment tumor and stroma regions of the tissue, segment cellular compartments, and phenotype infiltrating cells according to cell marker expression. Trained phenotyping algorithms developed in inForm for each marker were applied across the entire image set and data were compiled and summarized using Phenoptr Reports software. Summarized findings for intratumoral regions were visualized with boxplots and heatmaps. Wilcoxon tests were used to compare groups, with a two-sided statistical significance threshold of *p* < 0.05.

### Quantative Signaling Pathway Assay

All pre-BCG samples for Cohort A were used for the RT-qPCR quantitative signal transduction assay (OncoSignal®). Five out of 132 samples failed due to depleted RNA. Samples were checked for RNA integrity and possible PCR disturbing contaminants via performing control PCRs on a number of reference genes. Samples that passed incoming Quality Control were processed further and the activity of ER, AR, MAPK, PI3K, HH, Notch and TGFb pathways were measured. In short: 90μl Purified and DNase treated RNA of tumor tissue is mixed with One-step RT-qPCR reagents (SuperScript III Platinum One-Step qRT-PCR Kit (Thermo Fisher Scientific, cat.no. 11732088) and Nuclease-free water according to the manufacturer’s instructions. 25 μl of this mix was added to each well of a 96 wells OncoSignal Testing Plate after which the plate is sealed. Each well contains dried-down primers and probes to specifically amplify and detect one of the target genes. The PCR cycling and detection was performed in a Bio-Rad CFX96 (Touch) Real-Time PCR Detection System according to the following protocol: 30’ 50°C for reverse transcription followed by initial denaturation at 95°C for 5’ and 45 cycles of denaturation (15’’ 95°) and annealing (30’’ 60°C). The raw fluorescence data were exported from the Bio-Rad device and uploaded into the Pathway Activity software to calculate the 7 pathway activity scores for every sample. Finally, samples were labeled according to the generated molecular subtypes and visualized with the use of boxplots for all pathways. *P*-values are based on Wilcoxon testing. A logistic regression model was generated to identify BRS3 vs BRS1/2 patients. Sensitivity, specificity and the area under the curve were calculated using the pROC package [87].

## Data Availability

All data produced are available online at https://github.com/CostelloLab/BRSpred.

https://www.synapse.org/BRSpred

## Abbreviations

BC: bladder cancer
BCG: Bacillus Calmette-Guérin
BRS: BCG response subtypes
CIS: carcinoma in situ
ECM: extracellular matrix
EMT: epithelial-to-mesenchymal transition
HR-NMIBC: high-risk non-muscle invasive bladder cancer
HG: high-grade
HR: high-risk
HHR: very high-risk
LVI: lymphovascular invasion
MIBC: muscle invasive bladder cancer
NMIBC: non-muscle invasive bladder cancer
PFS: progression-free survival
re-TURBT: repeat transurethral resection of the bladder tumor
RFS: recurrence-free survival
RC: radical cystectomy
TAM: tumor associated macrophages
TME: tumor microenvironment
TURBT: transurethral resection of the bladder tumor
VH: variant histology

## Supplementary Materials

∘ Materials and methods
∘ Fig. S1. Consensus clustering results, Kaplan-Meier estimates of survival and gene signatures associated with BCG response subtypes (BRS) in the training and testing set in Cohort A.
∘ Fig. S2. Single nucleotide variant analysis for BCG response subtypes in Cohort A.
∘ Fig. S3. Heatmap of predicted BCG response subtypes in Cohort B.
∘ Fig. S4. Signatures and progression-free survival of BCG response subtypes in external cohorts.
∘ Fig. S5. BCG response subtype analyses in patients who had undergone a re-TURBT in Cohort A.
∘ Fig. S6. Heatmap of signature genes for BCG response subtypes, lumfFiinal gene set enrichment in BRS2 and regulon analysis. Result based on combination of Cohort A+B.
∘ Fig. S7. Nomogram to improve risk stratification in HR-NMIBC and OncoSignals® pathway results.
∘ Fig. S8. Kaplan-Meier estimates of Progression-Free Survival (PFS) based on currently published (N)MIBC molecular subtypes (Cohort A+B combined).
∘ Fig. S9. Gene fusions, gene set enrichment analysis and single-nucleotide variants in n=34 paired pre- and post-BCG tumors from n=34 HR-NMIBC patients.
∘ Table S1: Resources & clinical data
∘ Table S2: Transcriptomic SNVs results
∘ Table S3: Differentially expressed genes
∘ Table S4: Pathway & VIPER analyses
∘ Table S5: Immune deconvolution results
∘ Table S6: Paired pre- vs post-BCG analyses
∘ Table S7: Gene fusions in the paired sample analysis
∘ Data file S1: Normalized gene expression table BCG-naive Cohort A+B
∘ Data file S2: Raw counts Cohort A and Cohort B
∘ Data file S3: PAMR classifier coefficients

## Acknowledgements

FC de Jong expresses his deep gratitude to the Dutch Foundation “De Drie Lichten”, which supports medical research at the University of Colorado Anschutz Medical Campus, Aurora, CO, USA. The authors express their gratitude to Vebjørn Kvikstad, Jolien Mensink, Sébastien Rinaldetti and Joep de Jong for their help with patient data collection and analyses. This work utilized the Biostatistics and Bioinformatics Shared Resource and the Human Immune Monitoring Shared Resource supported by CA046934. Finally, we thank Lars Dyrskjøt for providing us with the raw count tables of the UROMOL21 dataset.

## Funding

This work was generously supported by MRACE Grant no.107477 (TZ), the Anschutz Foundation (JCC, DT), FICAN Cancer Researcher by the Finnish Cancer Institute (TDL), and in part from CA075115 (DT).

## Author Contributions

Conceptualization: FCJ, DTL, JCC, and TZ

Methodology: FCJ, TDL, DT, TZ and JCC

Formal analysis FCJ, TDL and JCC

Investigation: FCJ, TDL, RH and AM

Resources: RH, KJ, EJ, EB, DS, BN, EJ, JLB, DT, and TZ

Data curation: FCJ, TDL and AM

Writing—original draft preparation: FCJ

Writing—review and editing: FCJ, TM, JLB, DT, JCC and TZ

Supervision: DT, JCC and TZ

Project administration: JCC and TZ

Funding acquisition: DT, JCC and TZ

## Competing interests

JCC is co-founder of PrecisionProfile. None of the contributing authors have any competing interests, including specific financial interests and relationships and affiliations relevant to the subject matter in the manuscript.

## Data and materials availability

A detailed overview of all resources including assays, commercial systems and companies, software, critical R packages including versions and associated references used in this manuscript are in **Table S1**. The total clinical data table is included as **Table S1** as well. Raw data will be deposited for public use in the European Genome-Phenome Archive upon paper acceptance.

## Notes

### Competing Interest Statement

J.C.C is co-founder of PrecisionProfile. All other contributing authors have no conflicts of interest, including specific financial interests and relationships and affiliations relevant to the subject matter in the manuscript.

### Author Declarations

IRB approval was obtained from the Erasmus MC Medical Research Committee (MEC-2018-1097).

