## Supplementary Methods & Figures for "Non-muscle Invasive Bladder Cancer Molecular Subtypes Predict Differential Response to Intravesical Bacillus Calmette-Guérin"

### Supplementary Materials

#### Materials & Methods

*Transcriptome sequencing*

In Cohort A, total RNA was selected for polyadenylated mRNA using poly-T oligo-attached magnetic beads according to the manufacturer's protocol (Ribo-Zero Magnetic Kit, Epicentre Illumina). The selected mRNA was randomly fragmented by addition of the fragmentation buffer (Trushear, Covaris). The NEBNext Ultra library was used for library construction (NEBNext Ultra RNA Library Prep Kit, New England Biolabs). The first strand of cDNA was synthesized using a random hexamer primer and M-MuLV Reverse Transcriptase (RNase H), followed by a second strand cDNA synthesis using DNA Polymerase I and RNase H. Double-stranded cDNA was purified with beads (AMPure XP, Beckman Coulter) and remaining overhanging cDNA was converted into blunt ends via exonuclease/polymerase activities. After adenylating 3' DNA fragments, NEBNext Adapters were ligated to enable hybridization. The AMPure XP system was used a second time in order to select optimal cDNA fragments (150-200bp). Finally, the cDNA library was PCR-amplified and purified with AMPure XP beads for a third time. Constructed libraries were diluted to 1.5ng/μl based on quantitative result by Qubit 2.0. RT-qPCR was used to accurately quantify the library effective concentration (>2nM). RNA-seq libraries were pooled and sequencing performed to an estimated total output of 50 million paired-end 150bp reads per sample (HiSeq4000, Illumina). Image data files from Illumina were transformed to raw reads by CASAVA base calling and sequenced reads were stored as Fastq files. Raw reads containing adapters, undetectable bases or with a low-quality value were trimmed. Alignment to hg38 human reference genome was performed with STAR and mapped reads were stored as BAM files [1]. Transcripts were counted with HTSeq software [2]. Raw counts, FPKM and TPM values were used for downstream analyses. Normalized counts were used for calculation of differential expression, pathway and regulon analyses as detailed in the following sections. FPKM values was used as input in the classifyT1BC package, TPM values as input for immune deconvolution.

In Cohort B, there were several key differences. Briefly, the NEBNext Ultra II Directional RNA Library Prep Kit for Illumina was used to process the samples. rRNA was depleted from total RNA using the NEBNext rRNA depletion kit. After fragmentation of the rRNA reduced RNA, a cDNA synthesis was performed. This was used for ligation with the sequencing adapters and PCR amplification of the resulting product. Yield and quality after sample preparation was measured with the Fragment Analyzer. The size of the resulting products was consistent with the expected size distribution (a broad peak between 300-500 bp). This prepQC check was also performed with the Fragment Analyzer. Based on the molarity samples were pooled and sequenced. A concentration of 1.1 nM of DNA was used to perform paired-end 150bp reads sequencing with an estimated total output of 80 million reads per sample on a NovaSeq6000 (Illumina, control software NCS v1.7), with unique dual indexing and unique molecular identifiers according to standard operating procedures. Image analysis, base calling, and quality check was performed with the Illumina data analysis pipeline RTA3.4.4 and Bcl2fastq v2.20.

*Tissue specimens and RNA extraction*

Hematoxylin and eosin (HE) slides served as templates for FFPE blocks or blank slides. The uropathologist identified tumor areas with ≥80% tumor cells at the invasive front for macrodissection with sterile 1 mm biopsy punches. Total RNA was isolated with the High Pure FFPET isolation kits (Roche), according to the manufacturer's instructions. All samples in Cohort A passed multiple quality control (QC) steps: RNA was quantified with UV spectrophotometer (Nanodrop 260/280), RNA integrity was evaluated by RT-qPCR using housekeeping genes (*HMBS*, *HPRT1*, *ACTB*) and the Bioanalyzer 2100 (Agilent). Samples amplified by RT-qPCR that contained ≥500ng cDNA passed QC and were designated for next generation sequencing library construction. For Cohort B: all samples were analyzed with UV spectrophotometer and the Fragment

Analyzer (DNF-471); samples for which ≥ 200ng cDNA was available passed QC and were included for sequencing.

*Differential expression & pathway analysis*

Cohort A and B were combined based on matching HGNC gene names. Transcripts without information were removed from the total dataset. A batch effect coefficient was included into the *DESeq2* model, after which a variance-stabilizing transformation (VST) was applied to the transcripts and differential gene expression was calculated using the DESeq2 R package [3, 4]. For pathway analyses, *Limmas* Batch Effect removal was applied to the data, and together with the Hallmarks geneset (MSigDB v7.1) were used as input in Gene Set Enrichment Analysis (GSEA) [5, 6]. At least 1000 permutations were performed, phenotype was used as permutation type and gene sets with <15 or >500 genes were removed. For pathway analyses, we deemed the outcome as statistically significant at FDR<0.25. Secondly, the *R* package *GSVA* was used to calculate a single-sample normalized enrichment scores using GSEA Hallmarks [7]. Statistical analyses was done using the Kruskall-Wallis non-parametric test followed by a Bonferroni multiple testing correction. Finally, the webtool *gProfiler* was used to perform functional enrichment of KEGG and Reactome pathways [8]. Results of significantly enriched genes and pathways were visualized with boxplots, volcano plots and heatmaps (R package *ggplot2, pheatmap* and *Enhanced Volcano*) [8-11]. Apart from *FGFR3* expression, which is the actual expression and not a derivative, all other signatures in the heatmap of panel B in **Fig. 1** and **Fig. S3-S5** are the mean sum of expression of genes as shown in **Fig. S6A**.

*Immune cell deconvolution*

Gene expression data was used to quantify tumor purity and immune cell abundance using the ESTIMATE algorithm [12]. TPM values of the batch-corrected and combined dataset (Cohort A + B) was used as input for the *Immunedeconv* R package [13]. Three deconvolution algorithms using TPM-normalized data were run, including quanTIseq, EPIC, and CIBERSORT in absolute mode. Output tables with immune cell fractions per sample were visualized with barplots. Differences between subtypes was determined using the Wilcoxon rank sum test. The Bonferroni correction was applied to adjust *p*-values for multiple testing (statistical significance set at *p*.adj < 0.05).

*Regulon analyses*

VST normalized and batch-corrected gene expression data (Cohort A + B) was supplied to the VIPER *R* package [14]. Matrices for specific sample sets were used to estimate regulatory activity between groups. The regulon was taken from the pre-defined bladder cancer ARACNe gene network [15]. The regulon object, the bootstrapped (100 permutations) signature matrix and the null model (1000 permutations) generated the msVIPER object, which contained protein-activity regulatory network results and shadow-analyses to identify confounding regulators. Finally, significantly altered regulators and regulatory genes as described by TCGA, and others, were visualized using heatmaps [16, 17]. Statistical significance was set at FDR < 0.25.

*Pre-BCG vs post-BCG analyses*

We used the classifier to predict the BRS in *n*=44 post-BCG high-grade recurrences and compared differences in subtype distribution. Furthermore, we analyzed molecular differences before and after BCG therapy by selecting paired samples from patients with both pre- and post-BCG samples available (*n*=34). Differential gene expression analysis, GSEA Hallmarks, KEGG and Reactome pathway analyses, immune deconvolution, VIPER regulon analyses and SNVs analysis were done as previously described.

*Transcriptomic SNVs*

Samtools and Picard were used to align reads to the hg38 human reference genome, sort the reads according to the genome coordinates, followed by screening out of duplicated reads. GATK3 was used to carry out SNV and INDEL calling [18]. The resulting VCFs were used as input for ANNOVAR to annotate the SNP site, which includes dbSNP, gene types, the 1000-genome project and COSMIC v70 [19]. In our final analyses, we included only exonic and non-synonymous SNVs, due to the high likelihood of miscalls in FFPE-based transcriptomic data without a normal comparison.

*Fusion gene analyses*

Fusion genes were detected through STAR-Fusion and in-silico validated through FusionInspector to create an annotated lists of total gene fusions samples from Cohort A [20]. We only analyzed only gene fusions found in both tumor samples of a unique patient (i.e. pre-BCG tumor and post-BCG recurrence) to avoid false positive findings. We normalized the number of detected junction reads based on sequencing depth and included top fusions in Table S9. Ribosomal and mitochondrial-like fusions were removed from the total list. Confirmed gene fusions were depicted with circosplots.

### Supplemental Figures

**Fig. S1. Consensus clustering results, Kaplan-Meier estimates of survival and gene signatures associated with BCG response subtypes (BRS) in the training and testing set in Cohort A.**
**A:** Overall progression free survival (PFS) for Cohort A vs Cohort B; *p*-value by log-rank test. **B:** Consensus matrix for three clusters in *n*=99 primary high-risk NMIBC patients treated with BCG (training set). **C:** PFS for *n*=99 primary high-risk NMIBC patients treated with BCG and stratified according to BRSs (training cohort); *p*-values are pooled log-rank test. **D:** PFS for *n*=33 primary high-risk NMIBC patients treated with BCG stratified according to predicted BRS (testing cohort); *p*-values are pooled log-rank test. **E:** Boxplots comparing enrichment results for training cohort (*n*=99) and testing cohort (*n*=33) for selected GSEA Hallmarks. F: Results of consensus clustering in entire Cohort A with immune signature genes removed. From left to right: consensus matrices (*n*=3-5). CDF-plot, delta-area plot and cluster-consensus. **Abbreviations**: BCG = Bacillus Calmette-Guérin; CDF = cumulative distribution function; GSEA = gene set enrichment analysis; NMIBC = non-muscle invasive bladder cancer; PFS = Progression-Free Survival.

**Fig. S2. Single nucleotide variant analysis for BCG response subtypes in Cohort A.**
**A:** Enriched canonical pathways with non-synonymous exonic single-nucleotide variants grouped by BRS in *n*=132 pre-BCG, high-risk NMIBC patients and treated with BCG (Cohort A). **Abbreviations**: BCG = Bacillus Calmette-Guérin; BRS = BCG response subtypes; NMIBC = non-muscle invasive bladder cancer.

**Fig. S3. Heatmap of predicted BCG response subtypes in Cohort B.**
Heatmap of gene signatures based upon prediction of the BRS in *n*=151 pre-BCG, high-risk NMIBC patients (validation cohort B). Variables from top to bottom: 1) Progression to MIBC; 2) BCG responders vs BCG failure; 3) 68-gene *carcinoma in situ* (CIS) signature [21];4) UROMOL21 NMIBC subtypes [22]; 5) T1BC subtyper [23]; 6) TCGA MIBC subtypes [17]; 7) Consensus MIBC subtypes [23]; 8) UNC MIBC subtypes [24]; 9) MDA MIBC subtypes [25]; 10) Lund BC subtypes [26]; 11) Clinicopathological parameters associated to pre-BCG tumors; 12) BRS gene signatures based on mean gene expression of selected genes. Included genes are based on gene set enrichment and literature (details in methods). **Abbreviations**: BCG = Bacillus Calmette-Guérin; BRS = BCG response subtype; EAU = European Association of Urology; MDA = MD Anderson; (N)MIBC = (non-)muscle-invasive bladder cancer; TCGA = The Cancer Genome Atlas; UNC = University of North Carolina.

**Fig. S4. Signatures and progression-free survival of BCG response subtypes in independent cohorts. A:** Heatmap of gene signatures grouped according to predicted BRSs in patients from UROMOL21 that received at least a single instillation of BCG. Patients are sorted based on BRS and UROMOL21 subtypes. BRS3 tumors correspond to Class 2B (and Class 2A) tumors. BRS1 and BRS2 tumors correspond to less aggressive Class 1 and Class 3 tumors. **B:** The first Kaplan-Meier plot estimate shows *n*=161 BCG-treated patients grouped according to BRS. Right: PFS for *n*=66 T1 BCG-treated patients only. **C:** Heatmap of gene signatures grouped according to predicted BRS in the TCGA cohort. Patients are sorted based on BRS and TCGA subtypes. BRS3 mostly corresponds to basal-squamous and luminal infiltrated tumors. Luminal papillary tumors are associated with BRS1 and BRS2. **D:** Heatmap of gene signatures grouped according to predicted BRSs in the Chicago cohort. Patients are sorted on BRS and T1 subtypes. **Abbreviations**: BCG = Bacillus Calmette-Guérin; BRS = BCG response subtypes; (N)MIBC = (non-)muscle invasive bladder cancer; PFS = progression-free survival; TCGA = The Cancer Genome Atlas.

**Fig. S5. BCG response subtype analyses in patients who had undergone a re-TURBT from Cohort A.** From left to right: Consensus matrix, progression-free survival (PFS) and heatmap of gene signatures associated to BRS for *n*=3 clusters in *n*=91 primary, high-risk NMIBC patients with re-TURBT and treated with BCG (Cohort A). *P*-values are pooled log-rank test. **Abbreviations**: BCG = Bacillus Calmette-Guérin; BRS = BCG response subtypes; MIBC = muscle-invasive bladder cancer; NMIBC = non-muscle invasive bladder cancer.

**Fig. S6. Heatmap of signature genes for BCG response subtypes, luminal gene set enrichment in BRS2 and regulon analysis in Cohort A+B combined.**
**A:** Heatmap for *n*=283 pre-BCG, high-risk NMIBC samples. Included genes are the basis for signature gene plot as depicted in **Figure 1**. Tumors are sorted according to the BRS and as in **Figure 2**. **B:** Heatmap for 26 BC regulators as identified by TGCA and others using single-sample VIPER in *n*=283 pre-BCG, high-

risk NMIBC tumors grouped by BRS (columns). Hierarchical clustering between samples confirms the the existence of differential subtypes. **C:** Enrichment plots for a luminal bladder cancer signature and a luminal vs mesenchymal signature (GSEA) in BRS2 vs BRS1 and BRS3. Results indicate BRS2 has significantly more luminal pathway activity than both BRS1 and BRS3. **Abbreviations**: BCG = Bacillus Calmette-Guérin; BRS = BCG response subtypes; GSEA = gene set enrichment analysis; VIPER = Virtual Inference of Protein Activity by Enriched Regulon Analysis.

**Figure 7. Nomogram to improve risk stratification in HR-NMIBC and OncoSignals® pathway results**
**A:** Example of a nomogram predicting the risk of progressive disease with input parameters based on significant findings from Cohort A and B as presented in Fig. 4C. Of note: a nomogram should be designed in a prospectively collected cohort with a real-world risk of progressive disease. **B:** Boxplots demonstrating results of OncoSignals® RT-qPCR signal transduction assay of n=127 pre-BCG, high-risk NMIBC and treated with BCG grouped by BRS in Cohort A; p-values are Wilcoxon tests of BRS1/2 vs BRS3; *p.adj<10-2; **p.adj<10-3; ***p.adj<10-4 ***p.adj<10-5. **Abbreviations:** BCG = Bacillus Calmette-Guérin; BRS = BCG response subtypes. HR-NMIBC = high-risk non-muscle invasive bladder cancer

**Fig. S8. Kaplan-Meier estimates of Progression-Free Survival (PFS) based on currently published (non-)muscle invasive bladder cancer subtypes (Cohort A+Cohort B combined).**
**A:** PFS stratified according to the UROMOL21 NMIBC subtypes [22]; *p*-value is pooled log-rank test. **B:** PFS stratified according to the T1BC subtypes [23]; *p*-value is pooled log-rank test. **C:** PFS stratified according to the TCGA MIBC subtypes [17]; *p*-value is pooled log-rank test. **D:** PFS stratified according to the MDA MIBC subtypes [25]; p-value is pooled log-rank test. **E:** PFS stratified according to the UNC MIBC subtypes [24]; p-value is log-rank test. **F:** PFS stratified according to the Consensus MIBC subtypes [23]; *p*-value is pooled log-rank test. **G**: PFS stratified according to the Lund BC subtypes [26]; p-value is pooled log-rank test. **Abbreviations**: (N)MIBC = (non-)muscle-invasive bladder cancer.

**Fig. S9. Gene fusions, gene set enrichment analysis and single-nucleotide variants in *n*=34 paired pre- and post-BCG tumors from n=34 HR-NMIBC patients.**
**A:** Circosplot depicting gene fusions present in both pre-BCG and post-BCG sample within the same patient. Highlighted *FGFR3--TACC3* fusions were found in three individuals. **B:** Heatmap of top 50 GSEA hallmarks sorted on pre- vs post-BCG samples and BRSs. Signatures that contributed to the BRS3 subtype in pre-BCG samples, now also contributed to the BRS3 subtype in post-BCG tumors. Other subtypes (from top to bottom) include TCGA [17], Consensus MIBC, [23] UNC MIBC [24], MDA MIBC [25], Lund BC [26] and UROMOL21 NMIBC [22]. **C:** Enriched mutations in post-BCG tumors (non-synonymous exonic single-nucleotide variants); *n*=68 tumor samples from *n*=34 patients are sorted on patient and pre- vs post-BCG tumors. **Abbreviations**: BCG = Bacillus Calmette-Guérin; CIS = carcinoma in situ; EAU = European Association of Urology; MDA = MD Anderson; MIBC = muscle-invasive bladder cancer; NMIBC = non-muscle invasive bladder cancer; TCGA = The Cancer Genome Atlas; UNC = University of North Carolina.

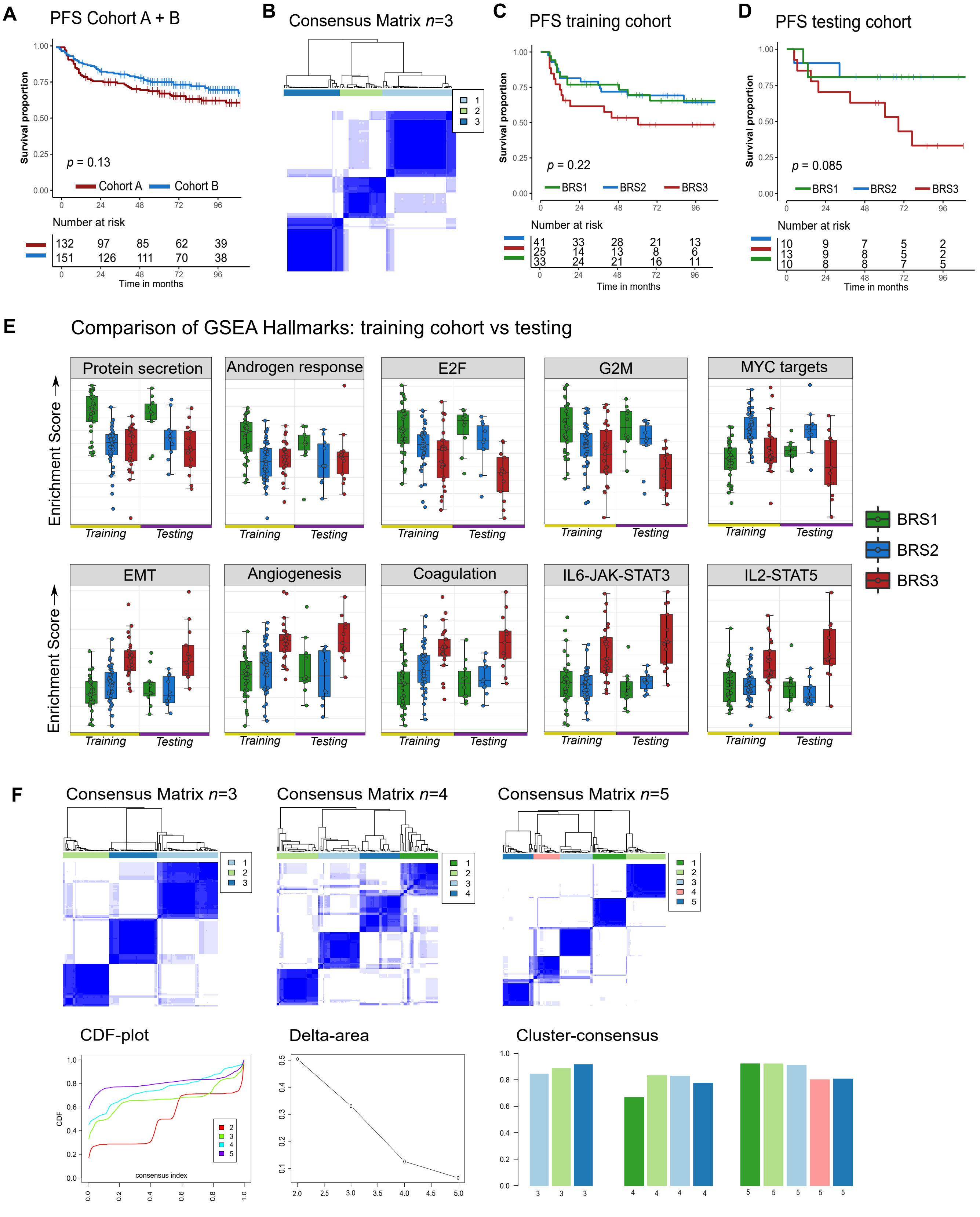


**Fig. S1. Consensus clustering results, Kaplan-Meier estimates of survival and gene signatures associated with BCG response subtypes (BRS) in the training and testing set in Cohort A.**

**A:** Overall progression free survival (PFS) for Cohort A vs Cohort B; p-value by log-rank test. **B:** Consensus matrix for three clusters in n=99 primary high-risk NMIBC patients treated with BCG (training set). **C:** PFS for n=99 primary high-risk NMIBC patients treated with BCG and stratified according to BRS (training cohort); p-values are pooled log-rank test. **D:** PFS for n=33 primary high-risk NMIBC patients treated with BCG stratified according to predicted BRS (testing cohort); p-values are pooled log-rank test. **E:** Boxplots comparing enrichment results for training cohort (n=99) and testing cohort (n=33) for selected GSEA Hallmarks. **F:** Results of consensus clustering in entire Cohort A with immune signature genes removed. From left to right: consensus matrices (n=3-5). CDF-plot, delta-area plot and cluster-consensus. **Abbreviations**: BCG = Bacillus Calmette-Guérin; CDF = cumulative distribution function; GSEA = gene set enrichment analysis; NMIBC = non-muscle invasive bladder cancer; PFS = Progression-Free Survival.

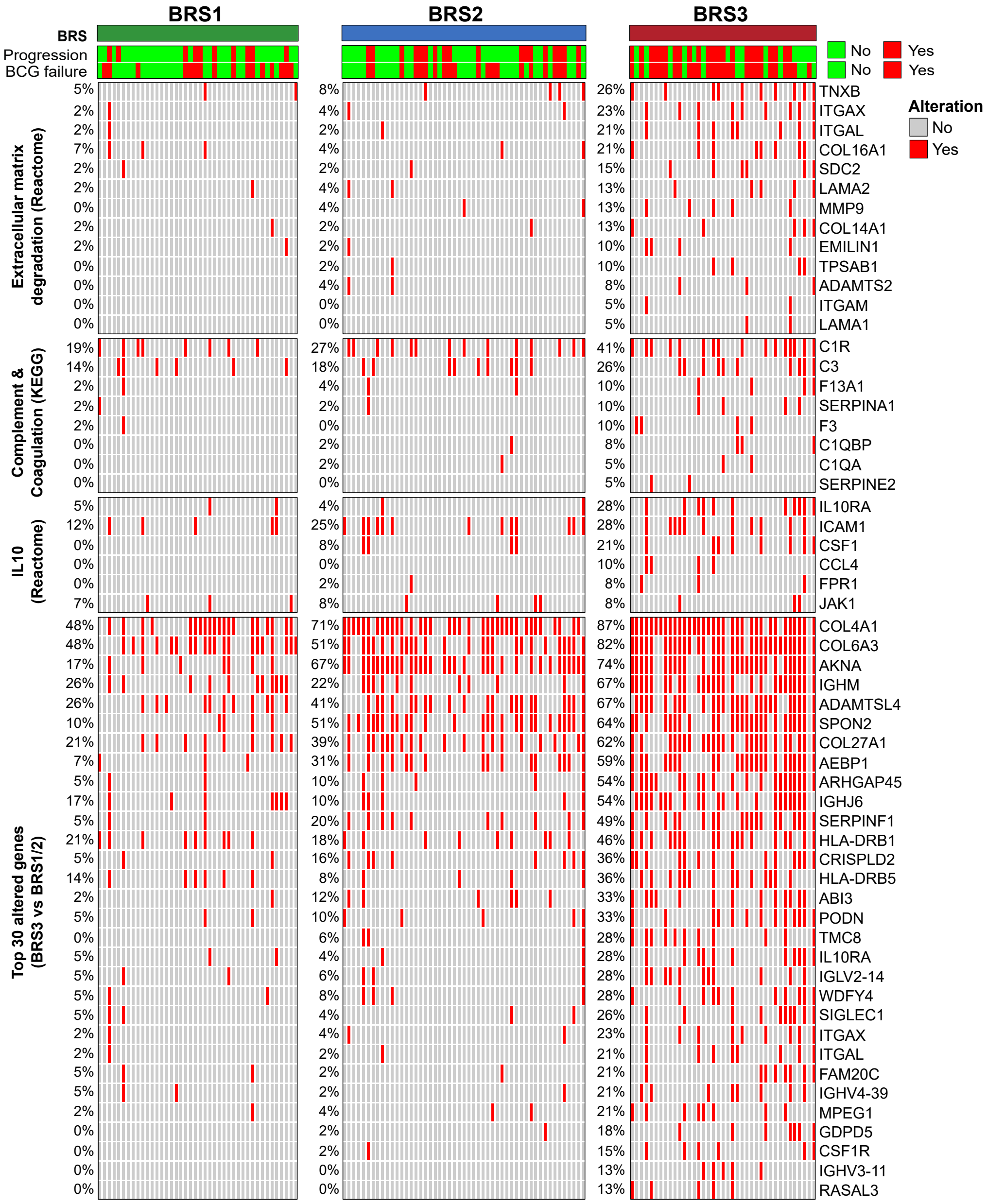


**Fig. S2. Single nucleotide variant analysis for BCG response subtypes in Cohort A.** Enriched canonical pathways with non-synonymous exonic single-nucleotide variants grouped by Erasmus-BRSs in n=132 pre-BCG, high-risk NMIBC patients and treated with BCG (Cohort A). Abbreviations: BCG = Bacillus Calmette-Guérin; BRS = BCG response subtypes; NMIBC = non-muscle invasive bladder cancer.

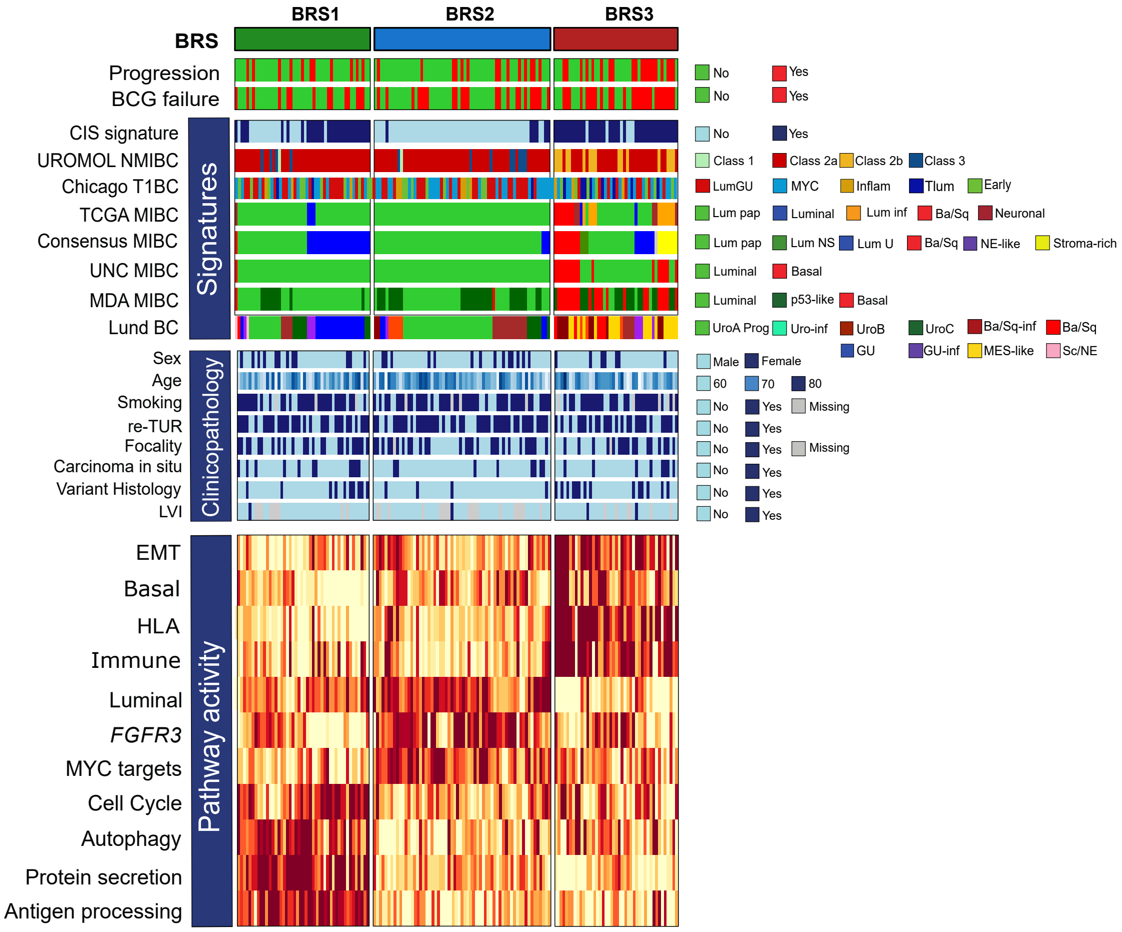


**Fig. S3. Heatmap of predicted BCG response subtypes in Cohort B.**
Heatmap of gene signatures based upon prediction of the BRS in n=151 pre-BCG, high-risk NMIBC patients (validation cohort B). Variables from top to bottom: 1) Progression to MIBC; 2) BCG responders vs BCG failure; 3) 68-gene carcinoma in situ (CIS) signature;4) UROMOL21 NMIBC subtypes; 5) T1BC subtyper; 6) TCGA MIBC subtypes; 7) Consensus MIBC subtypes; 8) UNC MIBC subtypes; 9) MDA MIBC subtypes; 10) Lund BC subtypes; 11) Clinicopathological parameters associated to pre-BCG tumors; 12) BRS gene signatures based on mean gene expression of selected genes. Included genes are based on gene set enrichment and literature (details in methods). **Abbreviations:** BCG = Bacillus Calmette-Guérin; BRS = BCG response subtype; EAU = European Association of Urology; MDA = MD Anderson; (N)MIBC = (non-)muscle-invasive bladder cancer; TCGA = The Cancer Genome Atlas; UNC = University of North Carolina**.**

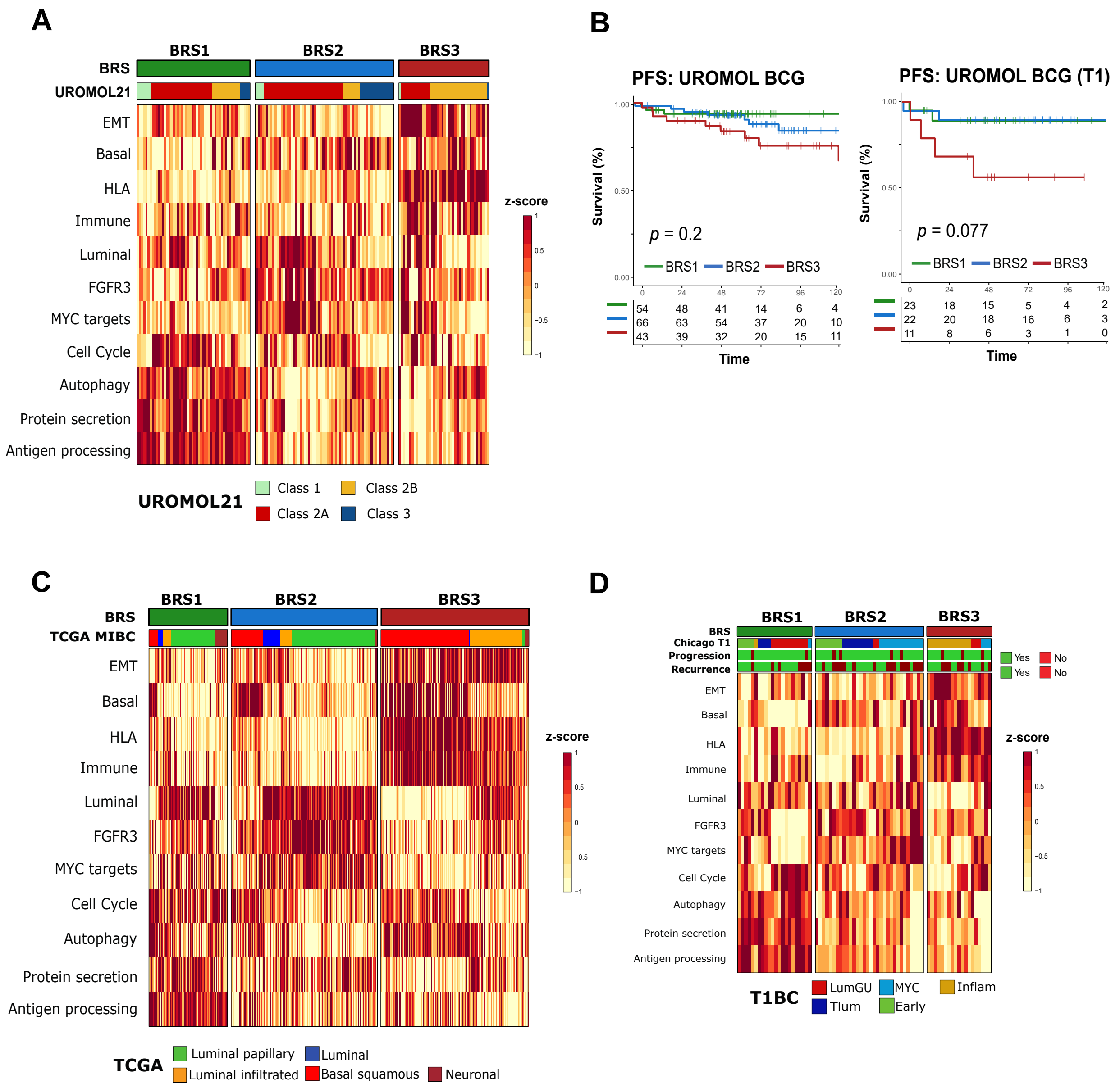


**Fig. S4. Signatures and progression-free survival of BCG response subtypes in external cohorts.**
**A:** Heatmap of gene signatures grouped according to predicted BRS in patients from UROMOL21 that received at least a single instillation of BCG. Patients are sorted based on BRS and UROMOL21 subtypes. BRS3 tumors correspond to Class 2B (and Class 2A) tumors. BRS1 and BRS2 tumors correspond to less aggressive Class 1 and Class 3 tumors. **B:** The first Kaplan-Meier plot estimate shows n=163 BCG-treated patients grouped according to BRS. Right: PFS for n=66 T1 patients that received BCG at least once. **C:** Heatmap of gene signatures grouped according to predicted BRS in the TCGA cohort. Patients are sorted based on BRS and TCGA subtypes. BRS3 mostly corresponds to basal-squamous and luminal infiltrated tumors. Luminal papillary tumors are associated with BRS1 and BRS2. **D:** Heatmap with T1BC subtypes. Gene signatures grouped according to predicted BRS in the T1BC cohort. Patients are sorted on BRS. **Abbreviations:** BCG = Bacillus Calmette-Guérin; BRS = BCG response subtypes; (N)MIBC = (non-)muscle invasive bladder cancer; PFS = progression-free survival; TCGA = The Cancer Genome Atlas.

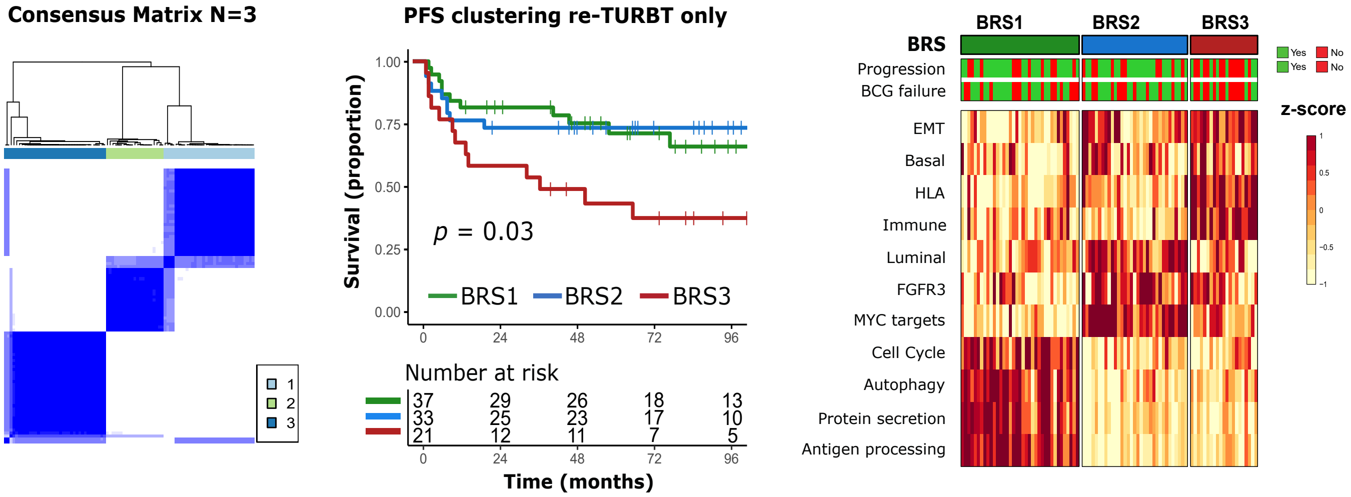


**Fig. S5. BCG response subtype analyses in patients who had undergone a re-TURBT from Cohort A.**
From left to right: Consensus matrix, progression-free survival (PFS) and heatmap of gene signatures associated to BRS for *n*=3 clusters in *n*=91 primary, high-risk NMIBC patients with re-TURBT and treated with BCG. P-values are pooled log-rank test. Abbreviations: BCG = Bacillus Calmette-Guérin; BRS = BCG response subtypes; MIBC = muscle-invasive bladder cancer; NMIBC = non-muscle invasive bladder cancer.

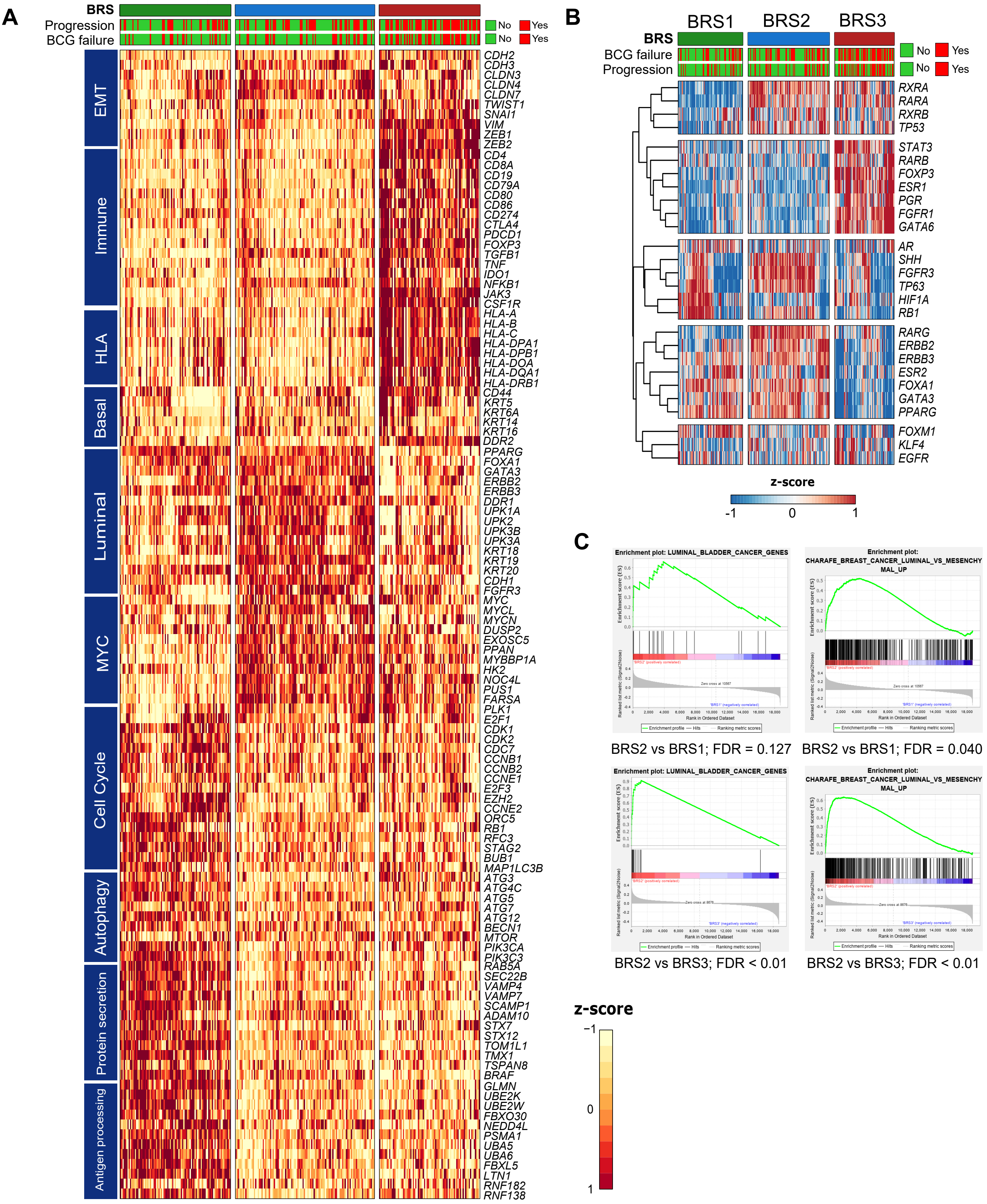


**Fig. S6. Heatmap of signature genes for BCG response subtypes, luminal gene set enrichment in BRS2 and regulon analysis. Result based on combination of Cohort A+B.**

**A:** Heatmap for n=283 pre-BCG, high-risk NMIBC samples. Included genes are the basis for signature gene plot as depicted in Figure 1. Tumors are sorted according to the BRS and as in Figure 2. **B:** Heatmap for 26 BC regulators as identified by TGCA and others using single-sample VIPER in n=283 pre-BCG, high- risk NMIBC tumors grouped by BRS (columns). Hierarchical clustering between samples confirms the the existence of differential subtypes. **C:** Enrichment plots for a luminal bladder cancer signature and a luminal vs mesenchymal signature (GSEA) in BRS2 vs BRS1 and BRS3. Results indicate BRS2 has significantly more luminal pathway activity than both BRS1 and BRS3. **Abbreviations:** BCG = Bacillus Calmette-Guérin; BRS = BCG response subtypes; GSEA = gene set enrichment analysis; VIPER = Virtual Inference of Protein Activity of Enriched Regulon Analysis.

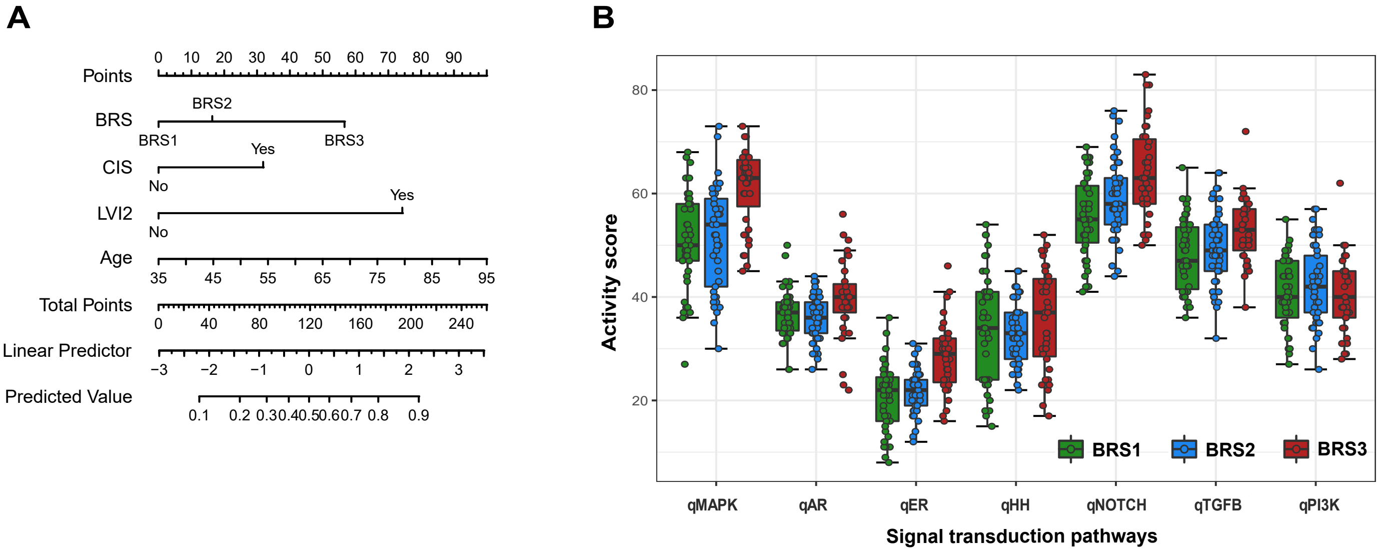


**Fig. S7. Nomogram to improve risk stratification in HR-NMIBC and OncoSignals® pathway results**

**A:** Example of a nomogram predicting the risk of progressive disease with input parameters based on significant findings from Cohort A and B as presented in Fig. 4C. Of note: a nomogram should be designed in a prospectively collected cohort with a real-world risk of progressive disease. B**:** Boxplots demonstrating results of OncoSignals® RT-qPCR signal transduction assay of n=127 pre-BCG, high-risk NMIBC and treated with BCG grouped by BRS in Cohort A; p-values are Wilcoxon tests of BRS1/2 vs BRS3; *p.adj<10-2; **p.adj<10-3; ***p.adj<10-4 ***p.adj<10-5. **Abbreviations:** BCG = Bacillus Calmette-Guérin; BRS = BCG response subtypes. HR-NMIBC = high-risk non-muscle invasive bladder cancer

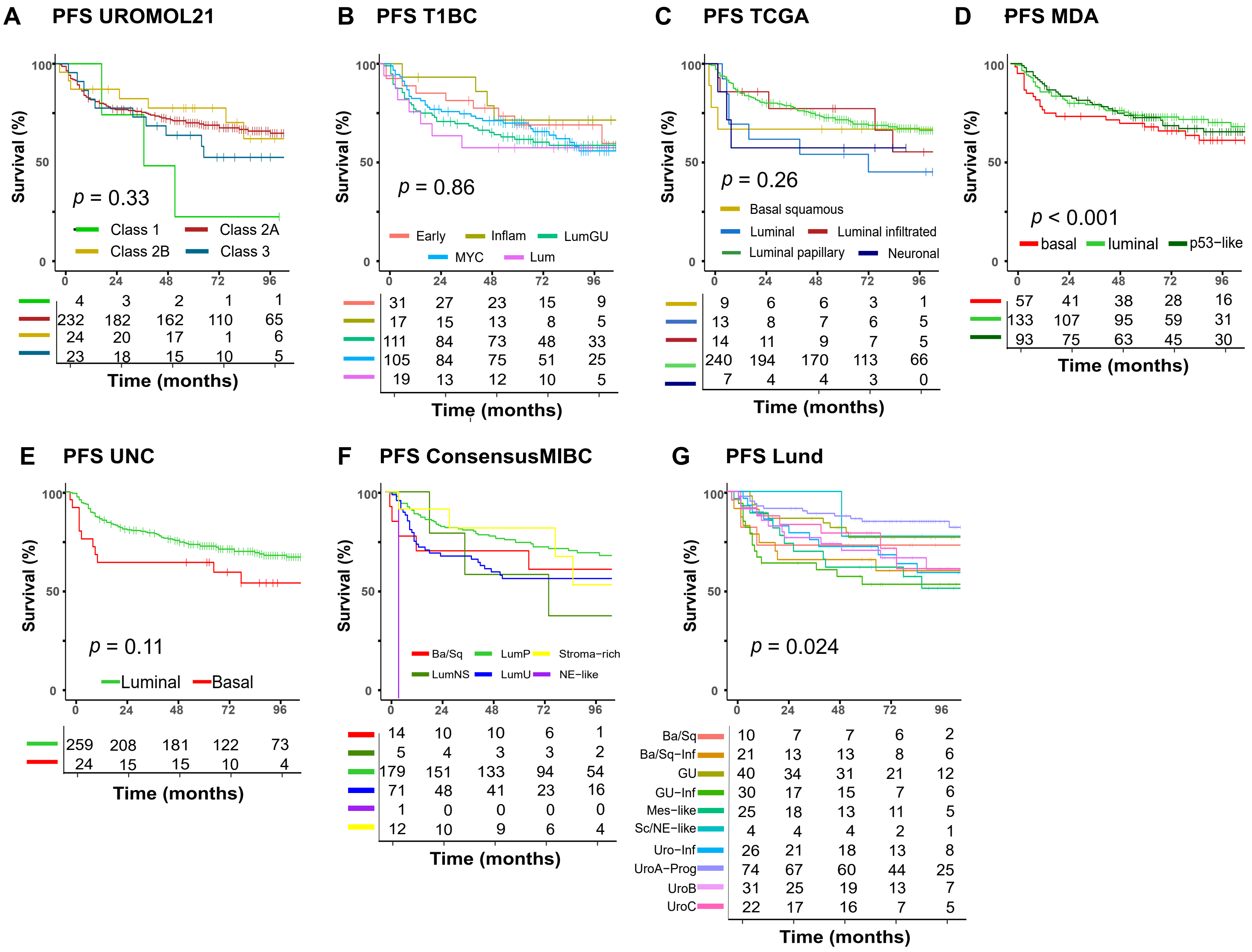


**Fig. S8. Kaplan-Meier estimates of Progression-Free Survival (PFS) based on currently published (N)MIBC molecular subtypes (Cohort A+B combined).**
**A:** PFS stratified according to the UROMOL21 NMIBC subtypes [22]; p-value is pooled log-rank test. **B:** PFS stratified according to the T1BC subtypes [23]; p-value is pooled log-rank test. **C:** PFS stratified according to the TCGA MIBC subtypes [17]; p-value is pooled log-rank test. **D:** PFS stratified according to the MDA MIBC subtypes [25]; p-value is pooled log-rank test. **E:** PFS stratified according to the UNC MIBC subtypes [24]; p-value is log-rank test. **F:** PFS stratified according to the Consensus MIBC subtypes [23]; p-value is pooled log-rank test. **G:** PFS stratified according to the Lund BC subtypes [26]; p-value is pooled log-rank test. Abbreviations: (N)MIBC = (non-)muscle-invasive bladder cancer.

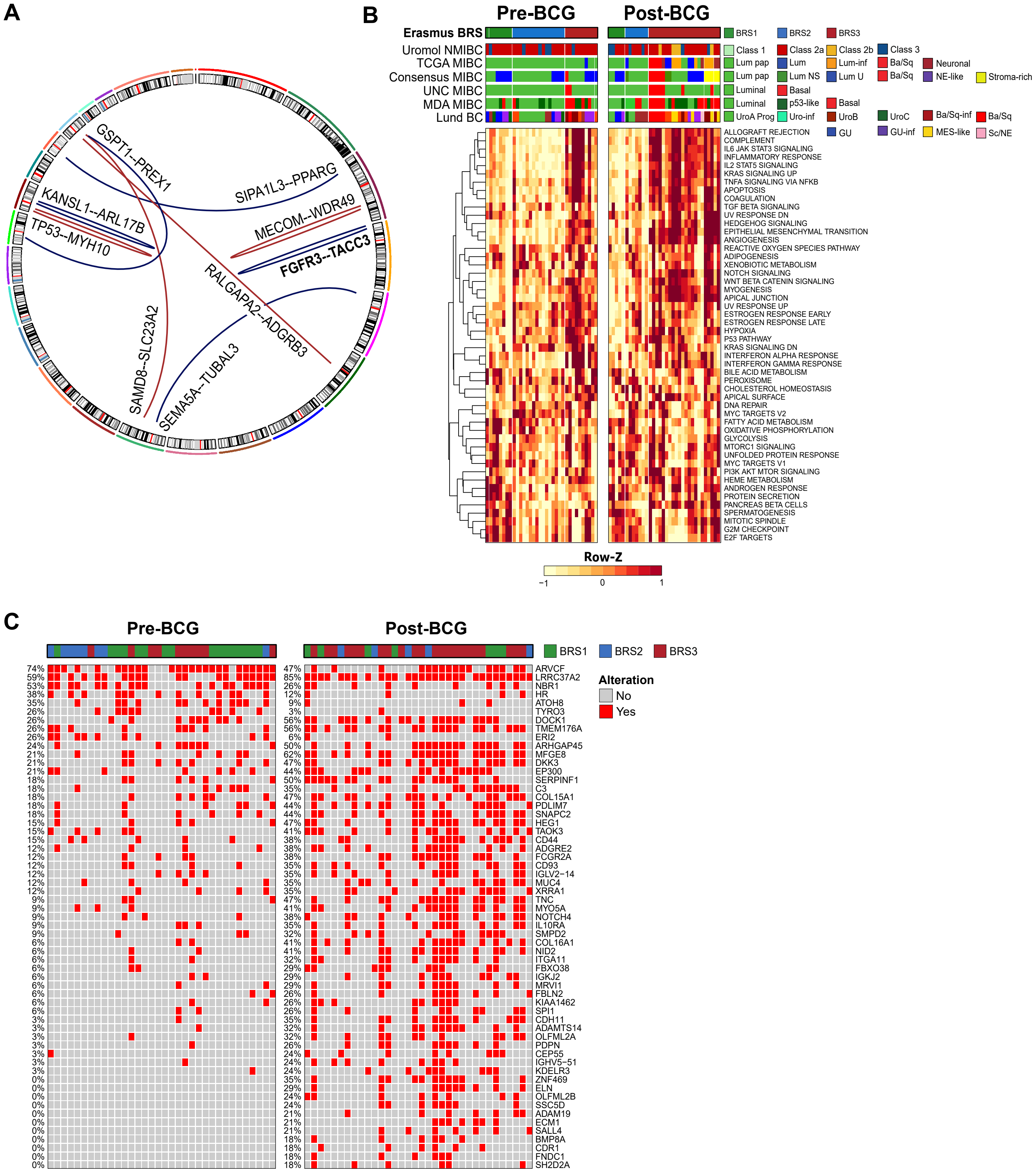


**Fig. S9. Gene fusions, gene set enrichment analysis and single-nucleotide variants in n=34 paired pre- and post-BCG tumors from n=34 HR-NMIBC patients.**

**A:** Circosplot depicting gene fusions present in both pre-BCG and post-BCG sample within the same patient. Highlighted *FGFR3--TACC3* fusions were found in three individuals. **B:** Heatmap of top 50 GSEA hallmarks sorted on pre- vs post-BCG samples and BRSs. Signatures that contributed to the BRS3 subtype in pre-BCG samples, now also contributed to the BRS3 subtype in post-BCG tumors. Other subtypes (from top to bottom) include TCGA (17), Consensus MIBC, (23) UNC MIBC (24), MDA MIBC (25), Lund BC (26) and UROMOL21 NMIBC (22). **C:** Enriched mutations in post-BCG tumors (non-synonymous exonic single-nucleotide variants); n=68 tumor samples from n=34 patients are sorted on patient and pre- vs post-BCG tumors. **Abbreviations:** BCG = Bacillus Calmette-Guérin; MDA = MD Anderson; (N)MIBC = (non-)muscle invasive bladder cancer; TCGA = The Cancer Genome Atlas; UNC = University of North Carolina.
